# Decreased hospitalizations and deaths from community-acquired pneumonia coincided with rising public awareness of personal precautions before the governmental containment and closure policy: A nationwide observational study in Japan

**DOI:** 10.1101/2022.06.23.22276838

**Authors:** Masato Tashiro, Shuntaro Sato, Akira Endo, Ryosuke Hamashima, Yuya Ito, Nobuyuki Ashizawa, Kazuaki Takeda, Naoki Iwanaga, Shotaro Ide, Ayumi Fujita, Takahiro Takazono, Kazuko Yamamoto, Takeshi Tanaka, Akitsugu Furumoto, Katsunori Yanagihara, Hiroshi Mukae, Kiyohide Fushimi, Koichi Izumikawa

**Author notes:** **Corresponding author:** Masato Tashiro, M.D., Ph.D., Department of Infectious Diseases, Nagasaki University Graduate School of Biomedical Sciences, 1-7-1 Sakamoto, Nagasaki 852-8501, Japan. Joint first authors (Masato Tashiro, Shuntaro Sato, Akira Endo).

## Abstract

**Background:** The effectiveness of population-wide compliance to personal precautions (mask-wearing and hand hygiene) in preventing community-acquired pneumonia has been unknown. In Japan, different types of non-pharmaceutical interventions from personal precautions to containment and closure policies (CACPs, e.g. stay-at-home requests) were sequentially introduced from late January to April 2020, allowing for separate analysis of the effects of personal precautions from other more stringent interventions. We quantified the reduction in community-acquired pneumonia cases and deaths and assessed if it coincided with the timing of increased public awareness of personal precautions before CACPs were implemented.

**Methods:** A quasi-experimental interrupted time series design was applied to non-COVID-19 pneumonia hospitalization and 30-day death data from April 2015 to August 2020 across Japan to identify any trend changes between February and April 2020. We also performed a comparative analysis of pyelonephritis and biliary tract infections to account for possible changes in the baseline medical attendance. These trend changes were then compared to multiple indicators of public awareness and behaviors related to personal precautions, including keyword usage in mass media coverage and sales of masks and hand hygiene products.

**Findings:** Hospitalizations and 30-day deaths from non-COVID-19 pneumonia dropped by 24.3% (95% CI 14.8 to 32.8, p < 0.001) and 16.1% (95% CI 5.5 to 25.5, p < 0.005) respectively in February 2020, before the implementation of CACPs, whereas pyelonephritis and biliary tract infections did not suggest a detectable change. These changes coincided with increases in indicators related to personal precautions rather than those related to contact behavior changes.

**Interpretation:** Community-acquired pneumonia could be reduced by population-wide compliance to moderate precautionary measures, such as wearing masks and hand hygiene.

**Funding:** JSPS KAKENHI Grant Number 22K17329 and JSPS Overseas Research Fellowships.

**Research in context:** *Evidence before this study:* The impact of personal precautions on community-acquired respiratory disease has been studied mainly for influenza and coronavirus infections, but no studies have evaluated the number of hospitalizations or 30-day deaths from overall non-COVID-19 pneumonia. We searched PubMed and medRxiv until May 3, 2022, for studies on the impact of personal precautions on community-acquired pneumonia using the following terms in the title and abstract: ((personal precaution*) OR (mask*) OR (non-pharmac*) OR (nonpharmac*)) AND (pneumonia). Before November 2019, when COVID-19 first emerged, no study was found that evaluated the impact of personal precautions such as masks on all-cause community-acquired pneumonia. After the emergence of COVID-19, there have been several reports of the relationship between non-pharmaceutical interventions (NPIs) and a decrease in non-COVID-19 pneumonia, but all reports evaluated the impact of general NPIs that aggregated different types of interventions, including personal precautions, physical distancing, and movement restrictions, and no studies were found that evaluated the impact on overall non-COVID-19 pneumonia from personal precautions alone.

*Added value of this study:* Our study found a reduction in hospitalizations and deaths from non-COVID-19 community-acquired pneumonia in Japan, especially those among the elderly population, had been detectable before the implementation of physical distancing policy and movement restrictions including stay-at-home requests. This reduction coincided with an increase in multiple indicators of public awareness of personal precautions, suggesting the potential benefit of population-level compliance to personal precautions (mask wearing and hand hygiene) against community-acquired pneumonia.

*Implications of all available evidence:* Maintaining a certain level of personal precautions in the population, e.g. by mask recommendations, may provide a positive public health impact even in the post-COVID era via reduced incidence of a spectrum of infectious diseases: most importantly, pneumonia as a major cause of death in the elderly. Since personal precautions are more sustainable than stringent restrictions such as lockdowns and could largely coexist with normal economic activities, long-term recommendations for personal precautions, at least in certain parts of the society, may warrant further discussion.

## Introduction

Community-acquired pneumonia is one of the world’s leading causes of death, with an estimated incidence of more than 300 million cases per year worldwide and a 30-day case fatality risk of over 10%.^1,2^ Since the emergence of the coronavirus disease 2019 (COVID-19) pandemic, a wide range of non-pharmaceutical interventions (NPIs) have been implemented to prevent its spread. In the meantime, it was reported that not only COVID-19 but also non-COVID-19 respiratory infections including community-acquired pneumonia showed a significant decline ^3^. Given the substantial disease burden from pneumonia, keeping a certain level of population-wide NPIs, if proven effective and sustainable, could be a new approach to the prevention of community-acquired pneumonia in society.

NPIs differ in their target, effectiveness, associated cost, and sustainability. Examples of NPIs that have been implemented to control the COVID-19 pandemic include universal masking, hand hygiene, physical distancing, and restrictions on people’s movement and gathering including so-called ‘containment and closure policies (CACPs)’.^4^ Some of these NPIs are not sustainable long-term and could only be implemented at the imminent threat of the outbreak, in particular CACPs. Here, we specifically refer to CACPs as restrictions on people’s social and economic activities that contribute to the domestic spread of infection, based on official orders/requests issued by the government, e.g., rules against large gatherings, physical distancing policy, lockdowns, closure of schools and public facilities, and regulations on non-essential businesses. On the other hand, personal precautions such as wearing masks and hand hygiene are less stringent measures that minimally interfere with social and economic activities and thus could be continued even after the COVID-19 pandemic, at least in certain settings, for prevention of community-acquired pneumonia. It should be noted, however, that even such moderate precautionary measures may not completely be free from detrimental aspects; for example, wearing masks has been reported to potentially impair facial recognition and cause skin problems.^5,6^ To discuss whether the potential benefits outweigh these disadvantages and to reach a social consensus on the post-pandemic ‘new normal’, the reduction in pneumonia incidence (and potentially that of other infectious diseases) associated with personal precautions needs to be quantified. During the COVID-19 pandemic, CACPs and personal precautions were mostly implemented in parallel and thus their separate effects on transmission dynamics are not necessarily clear. To elucidate the possible impact of population-wide personal precautions on pneumonia in the post-pandemic settings, assessment needs to focus on the effect of personal precautions in the absence of the impact of CACPs.

Experience of the COVID-19 epidemic in Japan may provide a useful case study to separate possible effects of personal precautions from those of CACPs because of its unique initial course of the outbreak, which made it one of the first countries with a heightened public awareness. In Japan, the first case of COVID-19 was reported on January 14, 2020,^7^ shortly after the World Health Organization (WHO) had been informed of pneumonia cases of unknown cause, later recognized as COVID-19, on December 31, 2019. Japanese mass media started to highlight the importance of personal precautions such as masks and hand washing in COVID-19 prevention in mid to late January,^8^ which was followed by soaring demand for masks.^9^ In early February 2020, the quarantine operation on a cruise ship Diamond Princess anchored at the Yokohama port in Japan experiencing a large COVID-19 outbreak, which eventually resulted in 712 cases and 14 deaths,^10^ drew wide media coverage throughout Japan. The number of confirmed COVID-19 cases was only up to 1–4 per day in January but began to exponentially increase from mid-February to mid-March 2020, shaping the ‘first wave’ with a peak in April 2020.^7^ In April 2020, the state of emergency against the COVID-19 pandemic was declared for the first time in Japan, and the number of cases showed a decrease in May 2020. The trend reversed upward again in July 2020 following the lift of the state of emergency and this ‘second wave’ peaked out in August 2020. Over the course of the outbreak, the governmental responses including public messaging and nonbinding requests have been delivered in multiple steps. Japan’s Ministry of Health, Labor and Welfare recommended on February 5, 2020 that the public take personal precautions such as cough etiquette (wearing a mask or covering nose and mouth when coughing) and washing hands.^11,12^ The Japanese government officially compiled the basic policy for COVID-19 responses on February 25, 2020.^13^ The public was encouraged to wash their hands, practice good cough etiquette, and refrain from leaving home when they have symptoms. Remote work and staggered work hours were recommended. The Japanese government has also implemented a nationwide school closure from March 2, 2020.^14^ Despite these actions, the increasing trend of COVID-19 cases continued, and the Japanese government declared a state of emergency for seven major prefectures (Saitama, Chiba, Tokyo, Kanagawa, Osaka, Hyogo, and Fukuoka) on April 7, 2020, which was extended to the entire country on April 16, 2020.^15^ After observing the decrease of COVID-19 patients by early May, the target prefectures of the state of emergency were gradually reduced and the state of emergency was lifted in all prefectures by May 25, 2020. The state of emergency consisted of requests to voluntary restraints of the public and businesses, e.g. reducing unnecessary travels and gatherings and curtailing business hours of restaurants and bars (‘stay-at-home request’).^16^ Although this declaration was not legally binding unlike other similar policies abroad, many businesses complied with the requests, shortening their business hours, and shifting to telecommuting. The human mobility index showed a marked reduction during the state of emergency.^17^

As such, the types and the level of NPIs against COVID-19 implemented in Japan varied between January and April 2020. Personal precautions such as masks and hand washing were recommended and public awareness rose in late January to February after the wide recognition of COVID-19 threat in Japan; a limited range of CACPs such as school closures began in March 2020; and more stringent CACPs, i.e. the declaration of a state of emergency were implemented in April 2020. Therefore, by examining the timing of the trend change in community-acquired pneumonia incidence in Japan, we can assess if there was a detectable impact of population-level personal precautions on community-acquired pneumonia before the introduction of CACPs.

This study aims to quantify the reduction in the community-acquired pneumonia incidence associated with the timing of increased public awareness of personal precautions in Japan. To this end, we applied the interrupted time series design to pneumonia hospitalization and death data in Japan over the past 5 years. To account for possible changes in the healthcare-seeking behavior due to the COVID-19 pandemic, the incidence of hospitalizations and deaths from pyelonephritis and biliary tract infections, which are endogenous infections unlikely linked to masking and hand hygiene, were also analyzed for comparison. Finally, we compared these trend changes to multiple indicators of public awareness and behaviors related to NPIs: keyword usage in the mass media coverage, sales data for masks and hand hygiene products, and community mobility data.

## Methods

### Study design

We used a quasi-experimental interrupted time-series (ITS) design to analyze the trend change in the number of hospitalizations and deaths from non-COVID-19 pneumonia in a nationwide administrative claims database from April 2015 to August 2020 to assess if any trend change in non-COVID-19 pneumonia incidence is associated with a step change in the public awareness and behavior related to personal precautions.^18^

### Data source of patients

We used the Japanese Diagnostic Procedure Combination (DPC) database,^19^ a large-scale hospitalization registration database routinely collected from over 1,000 participating medical institutions in Japan. This database contains patient billing information and summary data collected at the time of discharge from participating institutions. In this study, we used basic information from the database: age, sex, the primary cause of hospitalization and known comorbidities upon admission (based on the International Classification of Diseases, 10th Edition (ICD-10) codes), and outcome at discharge. By including only hospitalization with pneumonia as a primary cause, we limited our analysis to community-acquired pneumonia. The disease as the primary cause of hospitalization is determined at the time of discharge. The severity of pneumonia was recorded based on a scoring system called A-DROP (<u>A</u>ge, <u>D</u>ehydration, <u>R</u>espiratory failure, <u>O</u>rientation disturbance, and a low blood <u>P</u>ressure),^20^ which is a version of CURB-65 (<u>C</u>onfusion, <u>U</u>rea nitrogen, <u>R</u>espiratory rate, <u>B</u>lood pressure, <u>65</u> years of age and older) adapted to Japanese patients with pneumonia.

The need for informed consent was waived and the study protocol was approved by the Clinical Research Ethics Committee of Nagasaki University Hospital, Nagasaki, Japan, (Clinical Research Ethics Committee number 20122126).

### Patient selection

For consistency, only data from hospitals that were able to submit data for every month during the entire period were used in this study. We identified hospitalized patients aged 18 years and older with pneumonia (J10-J18, J69) except for pulmonary tuberculosis, pulmonary mycosis, pulmonary parasitosis, and COVID-19, with pyelonephritis (N10, N12), and with biliary tract infections (K800, K803, K804, K810, K830) from April 2015 to August 2020. Among pneumonia, three types of pneumonia: pneumococcal pneumonia, aspiration pneumonia, and influenza pneumonia were used in our subgroup analysis. *Mycoplasma pneumoniae* pneumonia, another major cause of community-acquired pneumonia, was not included in subgroup analyses because it is known to follow nonregular multi-annual epidemic cycles in Japan.^21^ If the same person was registered for the same disease in the same month multiple times, these records were considered as duplicates and merged into a unique record. The elderly were defined as those aged 65 years or older.

### Statistical analyses

This study used a segmented regression model based on an ITS design to identify change points and estimate the magnitude of change (called level change). Here we refer to the change point as the time point at which an abrupt change in the outcome is assumed to have occurred. Candidate time points are February, March, and April 2020. The segmented regression model consists of an indicator variable indicating before and after the change point, time elapsed since April 2015, time elapsed since the change point, and a Fourier-transformed term of the time point to adjust for seasonality of infection. Since an ITS design is considered to be unsuitable for autocorrelated data, we assessed autocorrelations and performed Breusch-Godfrey tests.^22,23^ These assessments were also performed for each subpopulation stratified by disease, age, and pneumonia type.

To identify change points that best describe the data, we compared models with change points in February, March, and April 2020 with models with no change (‘null’ model) using the Bayesian information criterion (BIC).^24^ We considered a difference of 2 in BIC as significant, i.e., we treated models with the best BIC and those within a difference of 2 in BIC as equally substantially supported.^25^ However, where these ‘best models’ included a null model, we took a conservative approach and concluded that the existence of a change point was not sufficiently supported by data.

For the selected models with a change point, the level change rates (%), 95% confidence intervals (95% CI), and p values were estimated using either an ordinary least squares linear regression with the log-transformed outcome or a Poisson regression, depending on the sample size. As an additional analysis, we performed a controlled interrupted time-series analysis using endogenous infections (i.e., pyelonephritis and biliary tract infections) as controls to explicitly adjust for the potential changes in the baseline hospitalization during the COVID-19 pandemic.^26^

All analyses were performed using R version 4.1.1 (R Core Team, R Foundation for Statistical Computing, Vienna, Austria). Our analytic code is available at https://github.com/ShuntaroS/pneumoniaITS_Japan.

### Time trends in indicators of personal precautions and physical contacts

To associate observed changes in pneumonia incidence with awareness and behavior changes in the Japanese population, we collected multiple indicators of public awareness and behavior related to personal precautions and physical contacts: the number of major newspaper articles with specific keywords,^27-29^ sales of mask and hand sanitizers, and Google community mobility data.^17^ As we are particularly interested in the effect of personal precautions separated from changes in contact behavior, we made a distinction between indicators related to personal precautions, e.g., wearing masks and hand hygiene, and those related to physical contacts, e.g., social distancing and staying at home. See appendix (pp 2-3) for further details.

### Role of the funding source

The funders had no role in the design of the study; in the collection, analyses, or interpretation of data; in the writing of the manuscript, or in the decision to publish the results.

## Results

### Background characteristics of patients

We analyzed a dataset of 644,164 patients hospitalized for pneumonia, pyelonephritis, and biliary tract infections from April 2015 to August 2020 from 297 hospitals across Japan via a monthly-collected administrative claims database (Table 1 and appendix, p 4). Our dataset included 415,712 patients with pneumonia (non-elderly: 45,552, elderly: 370,160), 77,924 patients with pyelonephritis (non-elderly: 17,534, elderly: 60,390), and 150,528 patients with biliary tract infections (non-elderly: 29,425, elderly: 121,103). Figure 1 shows the monthly number of hospitalizations by pneumonia type. In the first half of the year 2020, we observed a decrease in the number of pneumonia hospitalizations in both the non-elderly and the elderly compared to previous years. ‘Other pneumonia’, i.e., pneumonia cases not categorized as either pneumococcal pneumonia, aspiration pneumonia, or influenza pneumonia, accounted for more than half of all pneumonia cases, and the marked reduction in this category of pneumonia explained the reduction in overall pneumonia cases.

**Table 1.** Summary of background characteristics by disease from April 2015 to August 2020.

| Characteristics |  | Pneumonia<br>N = 415,712 | Pyelonephritis<br>N = 77,924 | Biliary tract<br>infections<br>N = 150,528 |
| --- | --- | --- | --- | --- |
| Sex | Male | 242,111 (58%) | 27,109 (35%) | 85,674 (57%) |
|  | Female | 173,601 (42%) | 50,815 (65%) | 64,854 (43%) |
| Age, years | Median (Q1, Q3) | 82 (74, 88) | 78 (66, 86) | 77 (67, 85) |
|  | 18 to 64 | 45,552 (11%) | 17,534 (23%) | 29,425 (20%) |
|  | 65 and over | 370,160 (89%) | 60,390 (77%) | 121,103 (80%) |
| Severity of Pneumonia | Mild | 34,200 (8.2%) | - | - |
|  | Moderate | 163,756 (39%) | - | - |
|  | Severe | 47,348 (11%) | - | - |
|  | Extremely severe | 19,928 (4.8%) | - | - |
|  | Unknown | 150,480 (36%) | - | - |
| Pneumonia type | Pneumococcal pneumonia | 19,709 (4.7%) | - | - |
|  | Aspiration pneumonia | 147,251 (35%) | - | - |
|  | Influenza pneumonia | 1,543 (0.4%) | - | - |
|  | Other pneumonia | 247,214 (60%) | - | - |

Detail summary for charlson comorbidity index category by disease can be found in appendix (p 4).

**Figure 1:**
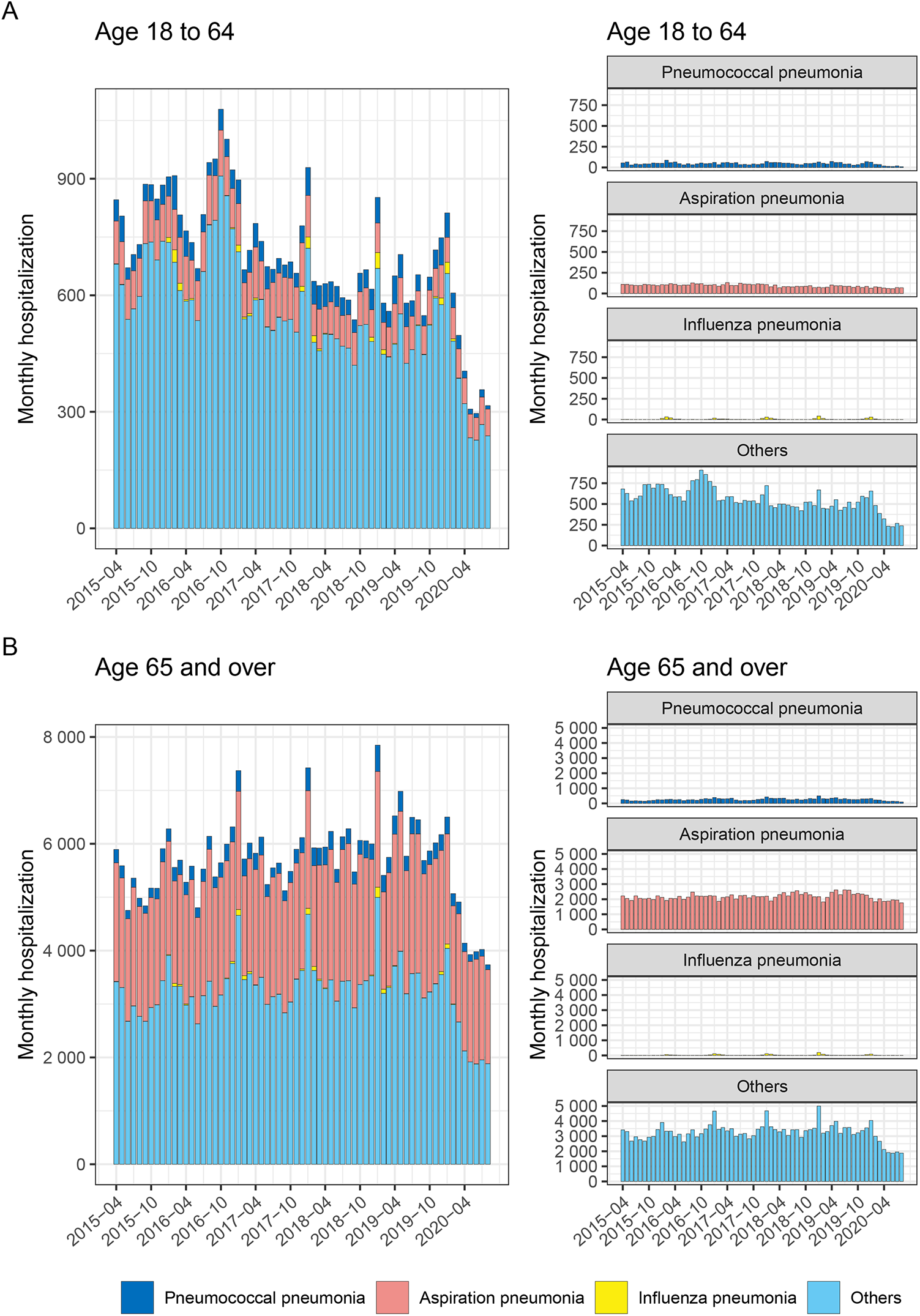
The number of monthly hospitalizations from community-acquired pneumonia among (A) non-elderly and (B) elderly adults by pneumonia type, April 2015 to August 2020, Japan. Colors represent different types of pneumonias; dark blue: pneumococcal pneumonia; orange: aspiration pneumonia; yellow: influenza pneumonia; and light blue: other pneumonia, which includes pneumonia of known (minor) causes and pneumonia of unknown/unclassified cause. Both stacked (left panels) and type-stratified (right panels) bar charts are shown.

### Model comparison

Our model comparison using BIC (Table 2) suggested that the model with a change point in February 2020 was almost consistently chosen as one of the best-supported models; in the few cases where the February model was unsupported, the model comparison did not reject the null model. Given these observations, we selected February 2020 as the change point of our primary interest and estimated the level change rates in February in the following analysis where applicable. The fitted and counterfactual curves in our ITS analysis can be found in the appendix (pp 5-12). Estimates obtained from models with other change points (March / April 2020) can be found in the appendix (pp 13-15) along with their BIC values.

**Table 2.**
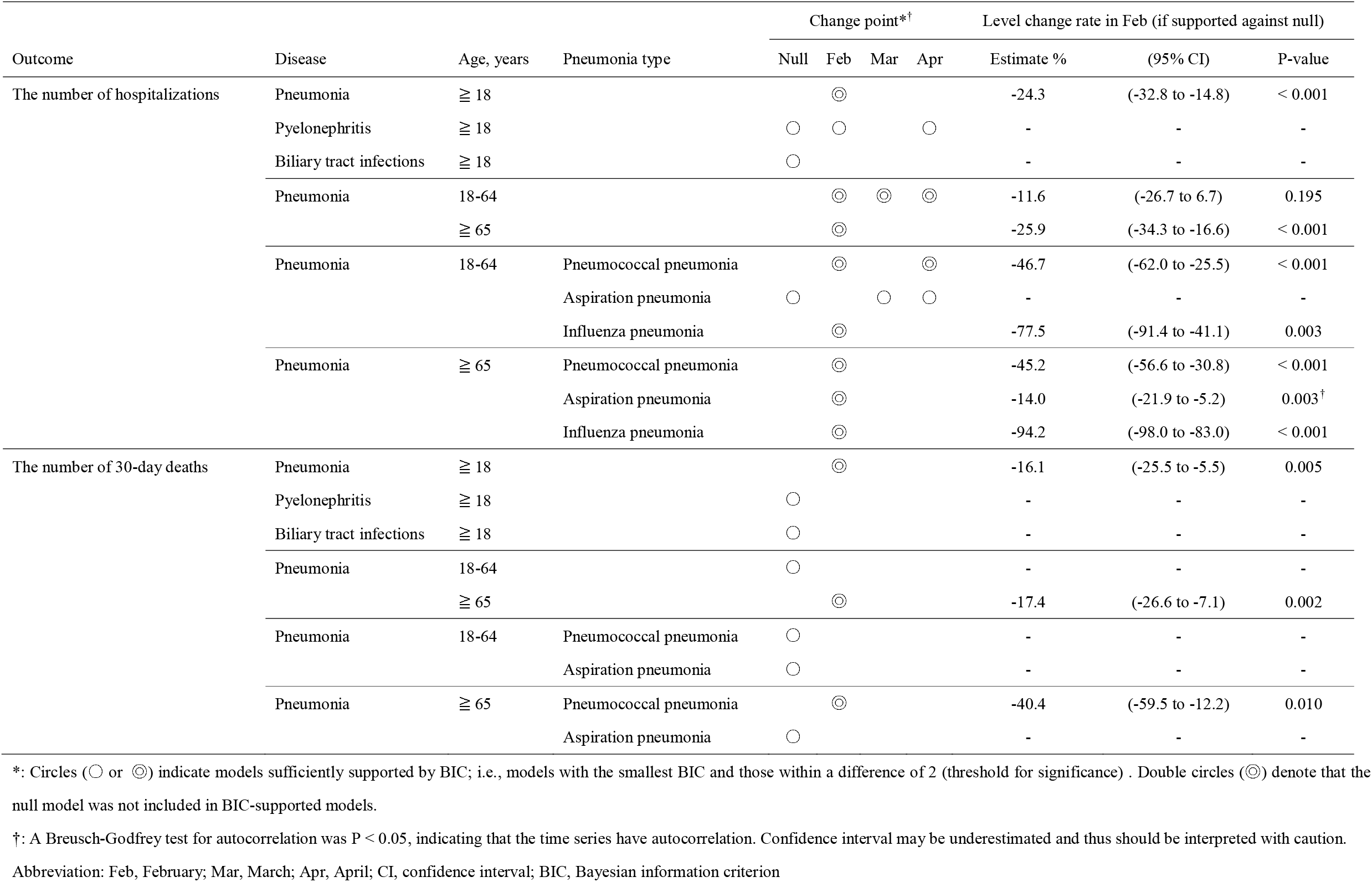
Estimated level change rate and change point for each outcome and sub-population in interrupted time-series analyses.

| Outcome | Disease | Age, years | Pneumonia type | Change point*<sup>†</sup> |  |  |  | Level change rate in Feb (if supported against null) |  |  |
| --- | --- | --- | --- | --- | --- | --- | --- | --- | --- | --- |
|  |  |  |  | Null | Feb | Mar | Apr | Estimate % | (95% CI) | P-value |
| The number of hospitalizations | Pneumonia | ≥ 18 |  |  | ⊙ |  |  | -24.3 | (-32.8 to -14.8) | < 0.001 |
|  | Pyelonephritis | ≥ 18 |  | ○ | ○ |  | ○ | - | - | - |
|  | Biliary tract infections | ≥ 18 |  | ○ |  |  |  | - | - | - |
|  | Pneumonia | 18-64 |  |  | ⊙ | ⊙ | ⊙ | -11.6 | (-26.7 to 6.7) | 0.195 |
|  |  | ≥ 65 |  |  | ⊙ |  |  | -25.9 | (-34.3 to -16.6) | < 0.001 |
|  | Pneumonia | 18-64 | Pneumococcal pneumonia |  | ⊙ |  | ⊙ | -46.7 | (-62.0 to -25.5) | < 0.001 |
|  |  |  | Aspiration pneumonia | ○ |  | ○ | ○ | - | - | - |
|  |  |  | Influenza pneumonia |  | ⊙ |  |  | -77.5 | (-91.4 to -41.1) | 0.003 |
|  | Pneumonia | ≥ 65 | Pneumococcal pneumonia |  | ⊙ |  |  | -45.2 | (-56.6 to -30.8) | < 0.001 |
|  |  |  | Aspiration pneumonia |  | ⊙ |  |  | -14.0 | (-21.9 to -5.2) | 0.003<sup>†</sup> |
|  |  |  | Influenza pneumonia |  | ⊙ |  |  | -94.2 | (-98.0 to -83.0) | < 0.001 |
| The number of 30-day deaths | Pneumonia | ≥ 18 |  |  | ⊙ |  |  | -16.1 | (-25.5 to -5.5) | 0.005 |
|  | Pyelonephritis | ≥ 18 |  | ○ |  |  |  | - | - | - |
|  | Biliary tract infections | ≥ 18 |  | ○ |  |  |  | - | - | - |
|  | Pneumonia | 18-64 |  | ○ |  |  |  | - | - | - |
|  |  | ≥ 65 |  |  | ⊙ |  |  | -17.4 | (-26.6 to -7.1) | 0.002 |
|  | Pneumonia | 18-64 | Pneumococcal pneumonia | ○ |  |  |  | - | - | - |
|  |  |  | Aspiration pneumonia | ○ |  |  |  | - | - | - |
|  | Pneumonia | ≥ 65 | Pneumococcal pneumonia |  | ⊙ |  |  | -40.4 | (-59.5 to -12.2) | 0.010 |
|  |  |  | Aspiration pneumonia | ○ |  |  |  | - | - | - |

\*: Circles (○ or ⊙) indicate models sufficiently supported by BIC; i.e., models with the smallest BIC and those within a difference of 2 (threshold for significance) . Double circles (⊙) denote that the null model was not included in BIC-supported models.
†: A Breusch-Godfrey test for autocorrelation was $P < 0.05$ , indicating that the time series have autocorrelation. Confidence interval may be underestimated and thus should be interpreted with caution.
Abbreviation: Feb, February; Mar, March; Apr, April; CI, confidence interval; BIC, Bayesian information criterion

### Changes in pneumonia and endogenous infections in adults

For the number of hospitalizations with pneumonia, the estimated level change rate in February was -24.3% (95% CI -32.8 to -14.8, p < 0.001) (Table 2). In contrast, we did not find sufficient support for a change in hospitalizations due to endogenous infections, i.e., pyelonephritis and biliary tract infections. In addition, the controlled interrupted time-series analysis using either of the endogenous infections as control also found a significant change in pneumonia hospitalizations in February (pyelonephritis as control: level change rate - 20.0%, 95% CI -30.6 to -7.7, p = 0.002; biliary tract infections as control: level change rate - 23.5%, 95% CI -34.0 to -11.3, p < 0.001). For the number of 30-day deaths with pneumonia, the estimated level change rate in February was -16.1% (95% CI -25.5 to -5.5, p < 0.005). We found no sufficient support for a level change in 30-day deaths in pyelonephritis and biliary tract infections.

### Changes in pneumonia by age group

The level change rates in February in the number of hospitalizations among non-elderly and elderly were estimated to be -11.6% (95% CI -26.7 to 6.7, p = 0.195) and -25.9% (95% CI - 34.3 to -16.6, p < 0.001), respectively (Table 2). The level change rate in February in the number of 30-day deaths with pneumonia for the elderly was estimated to be -17.4% (95% CI -26.6 to -7.1, p = 0.002), whereas no change in 30-day deaths with pneumonia was suggested for non-elderly.

### Changes in different types of pneumonia by age group

We also estimated the level change rates in the number of hospitalizations and deaths for different pneumonia types by age groups (Table 2). The number of hospitalizations showed significant reductions for all categories except for aspiration pneumonia among non-elderly. The number of 30-day deaths with pneumococcal pneumonia among the elderly showed a significant negative level change of -40.4% (95% CI -59.5 to 12.2, p = 0.010), while pneumococcal pneumonia in non-elderly and aspiration pneumonia in both non-elderly and elderly did not suggest a change. The 30-day deaths with influenza pneumonia did not allow for statistical analysis due to the small numbers observed.

### Trends in indicators related to personal precautions and physical contacts

We compared the temporal trend in the number of hospitalizations and 30-day deaths with pneumonia in adults with the trends in different indicators related to personal precautions and physical contacts in Japan (Figure 2). These indicators suggested that the changes in public awareness and behavior related to personal precautions (i.e., masks and hand hygiene) preceded those related to physical contacts by about 1–2 months. The number of articles in Japanese national newspapers with keywords related to personal precautions began to increase around January–February 2020 while the number of articles with keywords related to physical contacts did so in March 2020 onwards. Sales of masks and hand sanitizers showed a marked increase in January 2020. Google community mobility reports data (only available from mid-February 2020) showed its first drop of around 5% in March 2020, followed by a substantial drop of up to 30% in April onwards under the state of emergency. The observed change in pneumonia incidence in February 2020 thus coincided with changes in the indicators for personal precautions while the indicators for physical contacts showed marked changes only in March 2020 onward.

**Figure 2:**
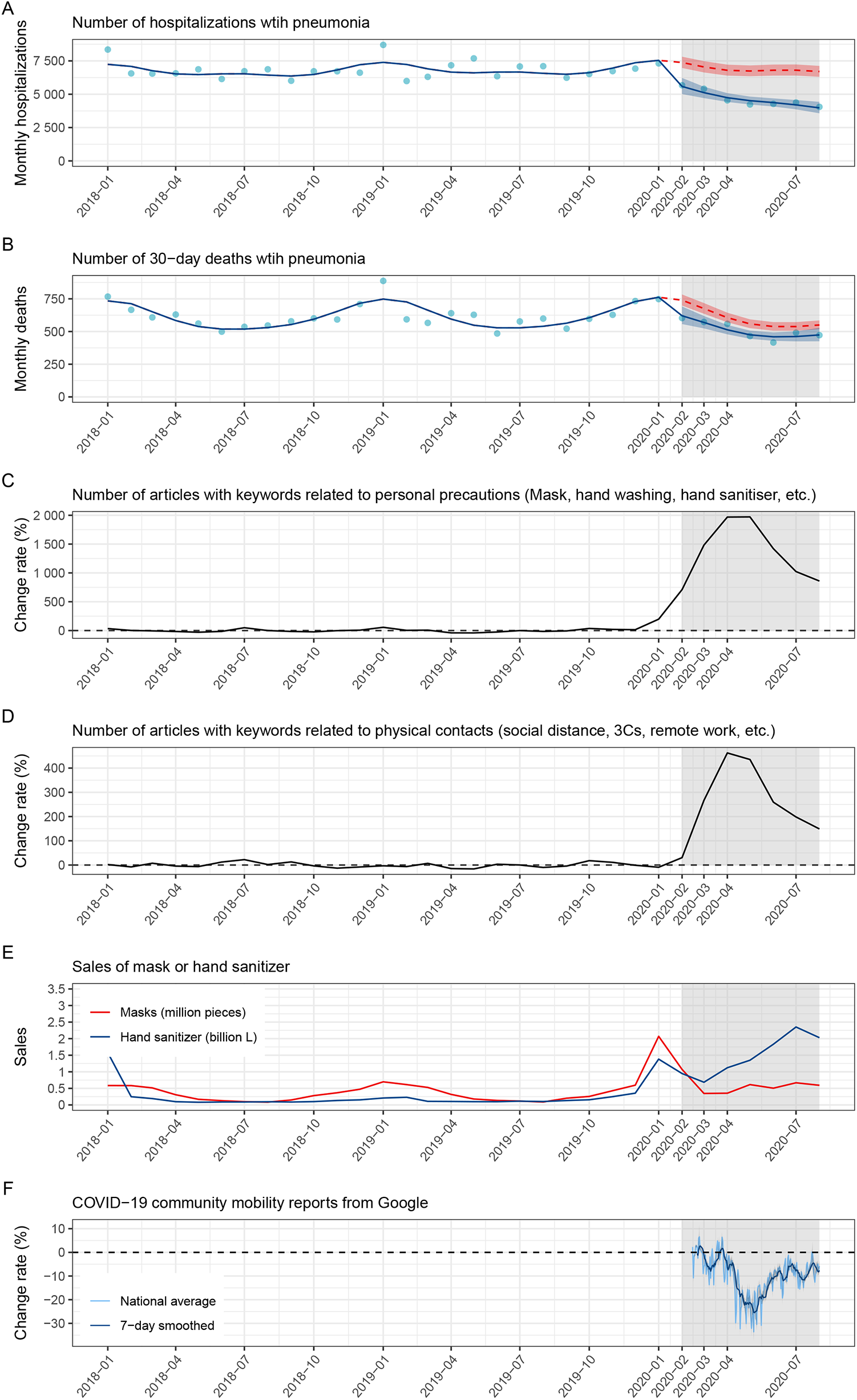
Interrupted time series analysis of the number of monthly (A) hospitalizations and (B) 30-day deaths from adult pneumonia and trends in indicators related to personal precautions and physical contacts (C, D: change rates in the monthly number of articles containing keywords related to personal precautions and physical contact; E: monthly sales of mask and hand sanitizer; F: change rate in Google community mobility data) for Japan between January 2018 and August 2020. Blue lines, red lines, and dots indicate the modeled curve fitted to the observed data, the counterfactual curve assuming no change point in February 2020, and the observed data, respectively. Ribbons indicate 95% confidence intervals. Gray-shaded areas indicate months at the change point (February 2020) and onward. The figures show data since January 2018 but estimates are based on data since April 2015. Change rates in the number of articles are relative to the average monthly number of articles between January 2018 and December 2019. Change rates in the Google community mobility data are relative to the day-of-the-week-wise average between January 3 and February 6, 2020, and is for non-residential location types (retail and recreation, grocery and pharmacy, parks, transit stations, and workplaces), presented in daily values and 7-day moving averages.

## Discussion

Using a quasi-experimental ITS design, we found a significant step reduction in non-COVID-19 adult pneumonia hospitalizations (−24.3%; 95% CI -32.8 to -14.8) and 30-day deaths (- 16.1%; 95% CI -25.5 to -5.5) in a Japanese national claims database in February 2020, when CACPs against COVID-19 had not yet been implemented in Japan. No detectable change was observed in the control time series of pyelonephritis and biliary tract infections, which suggests that the decrease in pneumonia incidence was not explained by a change in healthcare-seeking behavior. In our subgroup analyses, the temporal changes in February 2020 were generally clearer in among the elderly and in pneumonia with a known pathogen (pneumococcal and influenza pneumonia); e.g., the number of hospitalizations and deaths with pneumococcal pneumonia in the elderly both showed a drop of 45% and 40%, respectively in February 2020. The changes in hospitalizations and deaths among the non-elderly and in aspiration pneumonia deaths among the elderly were less clear. As the elderly cases accounted for 90% of pneumonia hospitalizations in the database, the observed drop in adult pneumonia hospitalizations and deaths in February 2020 was primarily driven by that in community-acquired pneumonia cases among the elderly. This drop in pneumonia observed in the elderly may be explained by limited community exposure to infectious agents under the population-level adherence to NPIs during the COVID-19 outbreak in Japan.

Our analysis of multiple indicators related to personal precautions (mask wearing and hand hygiene) and contact behaviors (physical contacts and community mobility) suggested that the reduction in community-acquired pneumonia incidence in February 2020 onward was most likely associated with elevated compliance to personal precautions. We found increases in the number of newspaper articles with keywords on personal precautions and in mask and hand sanitizer sales around January–February 2020. On the other hand, the number of articles with keywords on contact behavior changes, e.g., physical distancing and remote work, only showed a marked increase in March 2020 onward. Google community mobility data (available from mid-February 2020) also suggested that the change in people’s movement had been minimal at least until the end of February 2020, when the reduction in pneumonia incidence is already visible. These are in line with the fact that the Japanese government announced their first CACPs including recommendation of event cancellations and remote work in the final week of February,^13^ followed by more strong messages and measures such as school closures,^14^ recommendations of physical distancing, and declaration of the state of emergency in the following months.^15,16^

The dramatic voluntary change in personal precautions at the national level in the COVID-19 pandemic provided a historically valuable opportunity to assess the effectiveness of such population-wide adherence to personal precautions. Using an interrupted time-series design approach, we showed that decreased hospitalized patients and deaths from community-acquired pneumonia coincided with rising public awareness of personal precautions before the governmental CACPs. During the COVID-19 pandemic, a decrease in non-COVID-19 pneumonia has been reported worldwide,^3,30-33^ which was often attributed to the implementation of a wide range of NPIs including universal mask mandates, physical distancing policies, and lockdowns. However, the relative contributions of different types of NPIs are not necessarily well understood; the reduction in community-acquired pneumonia during the COVID-19 pandemic that has been reported to date was observed in the context of concurrent implementations of CACPs and personal precautions by the general public, and their separate effects could not be estimated. In this study, we attempted to exclude the influence of CACPs as well as the change in healthcare-seeking behavior or the capacity of the healthcare system due to the COVID-19 pandemic and assess the sole impact of personal precautions on the reduction of community-acquired pneumonia. Complementing existing studies on the effect of population-wide NPIs on pneumonia incidence, our study specifically suggested that a significant reduction in hospitalizations and deaths may even be achievable by personal precautions alone, e.g., mask wearing and hand hygiene, without restrictive CACPs. Compared to CACPs, personal precautions are less invasive and may be sustained at a certain level even after the COVID-19 pandemic. Our results may facilitate a discussion on whether the society accepts the continued population-wide adherence to personal precautions, at least in key public indoor settings, after the COVID-19 pandemic to mitigate the disease burden from community-acquired pneumonia.

The following limitations must be noted when interpreting the suggested causality between the decrease in community-acquired pneumonia in February 2020 and personal precautions such as masks and hand sanitizers in our ITS analysis. First, we could not obtain direct measures of compliance to personal precautions such as the proportion of people wearing masks or practicing hand hygiene around February 2020 and relied on surrogate indicators including mass media coverage and sales data. The earliest available data reported a mask-use rate of 60% in Japan already in the second week of March 2020,^34^ suggesting that at least mask usage had reached a substantially high level that could render a population-wide effect by early March. The unusual surge in mask demand in January 2020 is in line with an assumption that they were being used in February, when COVID-19 cases were constantly reported; however, we lack conclusive evidence on the actual mask usage to support this assumption. Second, possible changes in contact behavior that are not reflected in the community mobility index, e.g., voluntary event cancellations, were not assessed. However, the governmental request for event cancellation was announced on 25 February 2020 and an associated change in the growth rate of COVID-19 cases was only observed after this date;^35^ the impact of event cancellations on the population-level contact behavior in February 2020 was therefore arguably limited, if any. Third, although our model selection results were generally consistent with a choice of February 2020 as a change point (if there was a change), the possibility of a change in March or April was not completely excluded for some of the subanalyses, especially in the non-elderly with small case counts. We also did not consider multiple change points.

In conclusion, we showed a reduction in community-acquired pneumonia hospitalizations and deaths at the time of rising public awareness of personal precautions in Japan, before the government-led movement restrictions were commenced. Community-acquired pneumonia as a cause of death for many elderly people could be controlled if society as a whole continued to take moderate measures such as wearing masks and hand washing, which can be incorporated relatively easily into daily life compared to stringent and unsustainable restrictions.

## Supporting information

Appendix and ICMJE coi disclosure

## Data Availability

Epidemiological data on infectious diseases used in the analysis are available from the corresponding author upon reasonable request. Replication codes for the analysis are publicly available on GitHub (https://github.com/ShuntaroS/pneumoniaITS_Japan). The number of newspaper articles is available from the public databases cited. Sales data for masks and hand sanitizers were purchased from INTAGE Inc (Tokyo, Japan) and thus are not publicly available.

## Contributors

MT conceptualized the idea. SS and KF curated the data. SS and AE did data analysis. AE acquired funding. MT, SS, AE, and RH performed evidence collection. MT, SS, and AE developed the design methodology and creation of models. MT, A. Furumoto, K. Yanagihara, HM, and KI did management and coordination responsibility for the research activity planning and execution. MT, SS, and KF provided analysis data. SS did the programming. MT and KI supervised the planning and execution of research activities. MT, SS, AE, YI, NA, KT, NI, SI, A. Fujita, T. Takazono, K. Yamamoto, and T. Tanaka verified the data. MT, SS, and AE visualized the research results. MT, SS, and AE wrote the manuscript. All authors reviewed the manuscript and revised it critically.

## Declaration of interest

All authors declare any conflicts of interest using the ICMJE DISCLOSURE FORM.

## Acknowledgments

AE is supported by JSPS KAKENHI (22K17329) and JSPS Overseas Research Fellowships.

