## Appendix and ICMJE coi disclosure for "Decreased hospitalizations and deaths from community-acquired pneumonia coincided with rising public awareness of personal precautions before the governmental containment and closure policy: A nationwide observational study in Japan": Appendix_220624.pdf

*Joint first authors

[1]Department of Infectious Diseases, Nagasaki University Graduate School of Biomedical Sciences, Nagasaki, Japan; [2]Infection Control and Education Center, Nagasaki University Hospital, Nagasaki, Japan; [3]Clinical Research Center, Nagasaki University Hospital, Nagasaki, Japan; [4]Department of Infectious Disease Epidemiology, London School of Hygiene & Tropical Medicine, London, United Kingdom; [5]School of Tropical Medicine and Global Health, Nagasaki University, Nagasaki, Japan; [6]Department of Pulmonary Medicine, Graduate School of Medical Science, Kyoto Prefectural University of Medicine, Kyoto, Japan; [7]Department of Respiratory Medicine, Nagasaki University Hospital, Nagasaki, Japan; [8]Infectious Diseases Experts Training Center, Nagasaki University Hospital, Nagasaki, Japan; [9]Department of Laboratory Medicine, Nagasaki University Hospital, Nagasaki, Japan; [10]Department of Health Policy and Informatics, Graduate School of Medical and Dental Sciences, Tokyo Medical and Dental University, Tokyo, Japan.

**Corresponding author:**

Masato Tashiro, M.D., Ph.D., Department of Infectious Diseases, Nagasaki University Graduate School of Biomedical Sciences, 1-7-1 Sakamoto, Nagasaki 852-8501, Japan

### SUPPLEMENTAL MATERIAL

### SUPPLEMENTAL METHODS

*Time trends in indicators of personal precautions and physical contacts*

We collected newspaper article counts, mask and hand sanitizer sales data, and Google community mobility data to assess changes in public awareness and behaviors in the population in early 2020. As a measure of public awareness, we obtained the number of articles in two Japanese national newspapers containing specific keywords from January 2018 to August 2020 from their article databases.[1,2] Search keywords related to personal precautions included "masks" and "hand sanitizer" and those related to physical contact included "social distance", "3Cs", and "remote work".[3] All Japanese keywords used in the search and their English translations can be found in the below table. Using the average monthly number of articles between January 2018 and December 2019 as a baseline, we plotted the relative change in the number of articles in Figure 2 in the main text.

| | Original search formula in Japanese | English translations* |
|---|---|---|
| Personal precautions | "マスク" OR "咳エチケット" OR "手洗い" OR "石鹸" OR "石けん" OR "手指消毒" OR "手指衛生" | “masks” OR “cough etiquette” OR “hand washing” OR “soap” OR “hand sanitizer” OR “hand hygiene” |
| Physical contacts | "ソーシャルディスタンス" OR "社会的距離" OR "身体的距離" OR "3 密" OR "三密" OR "自粛" OR "自宅" OR "リモートワーク" OR "テレワーク" OR "オンライン会議" OR "時差通勤" OR "自宅勤務" OR "在宅勤務" OR "遠隔勤務" OR "休校" OR "学校休業" | “social distance” OR “physical distance” OR “3Cs” OR “voluntary restraint” OR “home” OR “remote work” OR “telework” OR “online meeting” OR “staggered commuting” OR “work at home” OR “school closure” |

* Duplicates are removed where multiple Japanese search terms translate into the same English.

As measures of observed behaviors, we also collected mask and hand sanitizer sales data (personal precautions) and Google community mobility data (physical contacts). Sales data was retrieved from a database provided by INTAGE Inc (Tokyo, Japan) from January 2018 through August 2020. Google community mobility data provides regional-level movement trends by location type (retail and recreation, grocery and pharmacy, parks, transit stations, workplaces, and residential) as a relative change from the day-of-the-week-wise average from January 3 through February 6, 2020. We obtained the daily national average of Google mobility in Japan in all location types but residential to measure the contact behavior of individuals outside the home.

**SUPPLEMENTAL TABLE 1. A detailed summary of Charlson comorbidity index categories**

| Charlson comorbidity Index | Pneumonia N = 415,712 | Pyelonephritis N = 77,924 | Biliary tract infections N = 150,528 |
|---|---|---|---|
| Myocardial Infarction | 7,600 (1·8%) | 1,079 (1·4%) | 2,817 (1·9%) |
| Congestive Heart Failure | 75,161 (18%) | 5,821 (7·5%) | 8,846 (5·9%) |
| Peripheral Vascular Disease | 7,645 (1·8%) | 1,112 (1·4%) | 2,141 (1·4%) |
| Cerebrovascular Disease | 65,710 (16%) | 9,104 (12%) | 12,926 (8·6%) |
| Dementia | 74,991 (18%) | 10,012 (13%) | 11,779 (7·8%) |
| Chronic Pulmonary Disease | 74,804 (18%) | 3,164 (4·1%) | 5,699 (3·8%) |
| Connective Tissue Disease-Rheumatic Disease | 12,791 (3·1%) | 2,042 (2·6%) | 1,710 (1·1%) |
| Peptic Ulcer Disease | 13,900 (3·3%) | 2,361 (3·0%) | 9,247 (6·1%) |
| Mild Liver Disease | 11,746 (2·8%) | 2,498 (3·2%) | 10,159 (6·7%) |
| Diabetes without complications | 60,814 (15%) | 12,936 (17%) | 26,381 (18%) |
| Diabetes with complications | 14,374 (3·5%) | 3,697 (4·7%) | 4,114 (2·7%) |
| Paraplegia and Hemiplegia | 1,724 (0·4%) | 350 (0·4%) | 308 (0·2%) |
| Renal Disease | 24,401 (5·9%) | 4,709 (6·0%) | 5,193 (3·4%) |
| Cancer | 47,295 (11%) | 9,467 (12%) | 28,344 (19%) |
| Moderate or Severe Liver Disease | 535 (0·1%) | 114 (0·1%) | 851 (0·6%) |
| Metastatic Carcinoma | 8,286 (2·0%) | 1,486 (1·9%) | 4,503 (3·0%) |
| AIDS/HIV | 139 (< 0·1%) | 17 (< 0·1%) | 15 (< 0·1%) |

**SUPPLEMENTAL FIGURE 1. Interrupted time series subanalyses of the number of monthly hospitalizations and 30-day deaths in Japan between April 2015 and August 2020.**

Blue lines, red lines, and dots indicate the modeled curve fitted to the observed data, the counterfactual curve assuming no change point in February 2020, and the observed data, respectively. Ribbons indicate 95% confidence intervals. Gray-shaded areas indicate months at the change point (February 2020) and onward. The figures show data from January 2018 but estimates are based on data from April 2015. The mapping between figures and details of the subanalyses is shown in the table below.

| Figure label | Outcome | Disease | Age, years | Pneumonia type |
|---|---|---|---|---|
| A | The number of hospitalizations | Pneumonia | ≧ 18 | |
| B | | Pyelonephritis | ≧ 18 | |
| C | | Biliary tract infections | ≧ 18 | |
| D | | Pneumonia | 18-64 | |
| E | | | ≧ 65 | |
| F | | Pneumonia | 18-64 | Pneumococcal pneumonia |
| G | | | | Aspiration pneumonia |
| H | | | | Influenza pneumonia |
| I | | Pneumonia | ≧ 65 | Pneumococcal pneumonia |
| J | | | | Aspiration pneumonia |
| K | | | | Influenza pneumonia |
| L | The number of 30-day deaths | Pneumonia | ≧ 18 | |
| M | | Pyelonephritis | ≧ 18 | |
| N | | Biliary tract infections | ≧ 18 | |
| O | | Pneumonia | 18-64 | |
| P | | | ≧ 65 | |
| Q | | Pneumonia | 18-64 | Pneumococcal pneumonia |
| R | | | | Aspiration pneumonia |
| S | | Pneumonia | ≧ 65 | Pneumococcal pneumonia |
| T | | | | Aspiration pneumonia |

**Outcome: The number of hospitalizations**

A

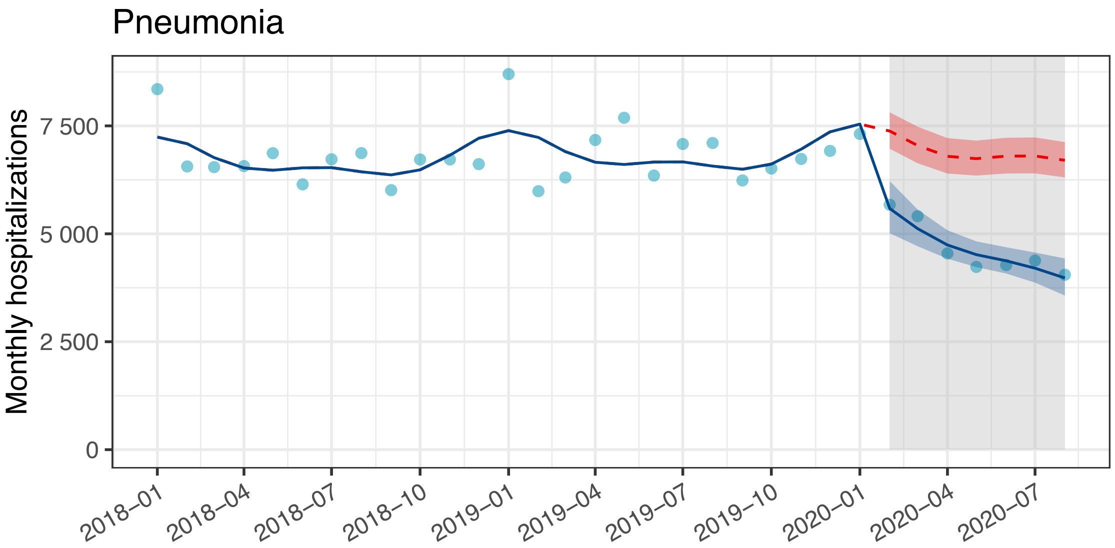


B

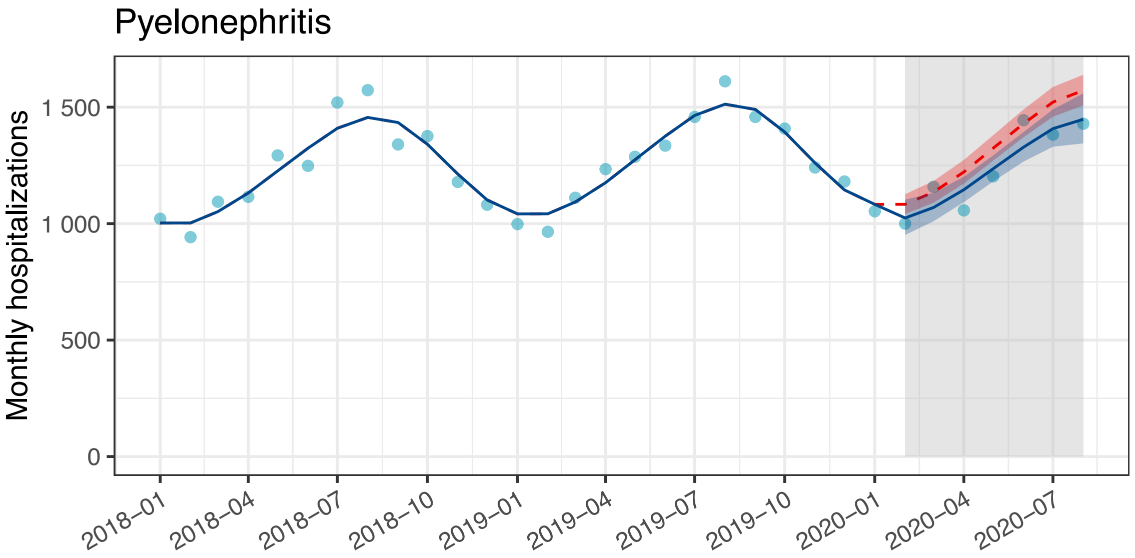


C

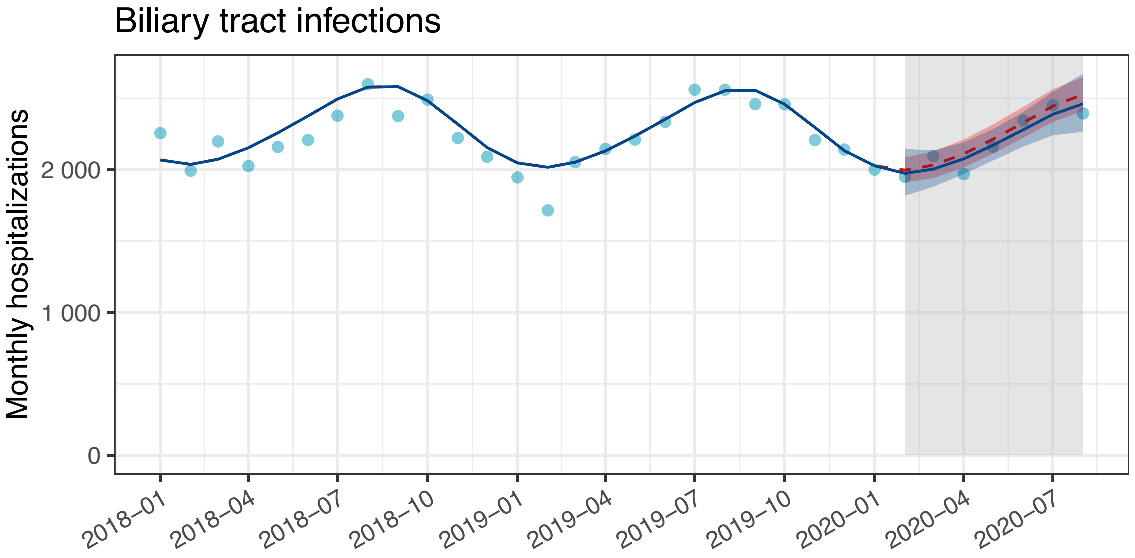

D

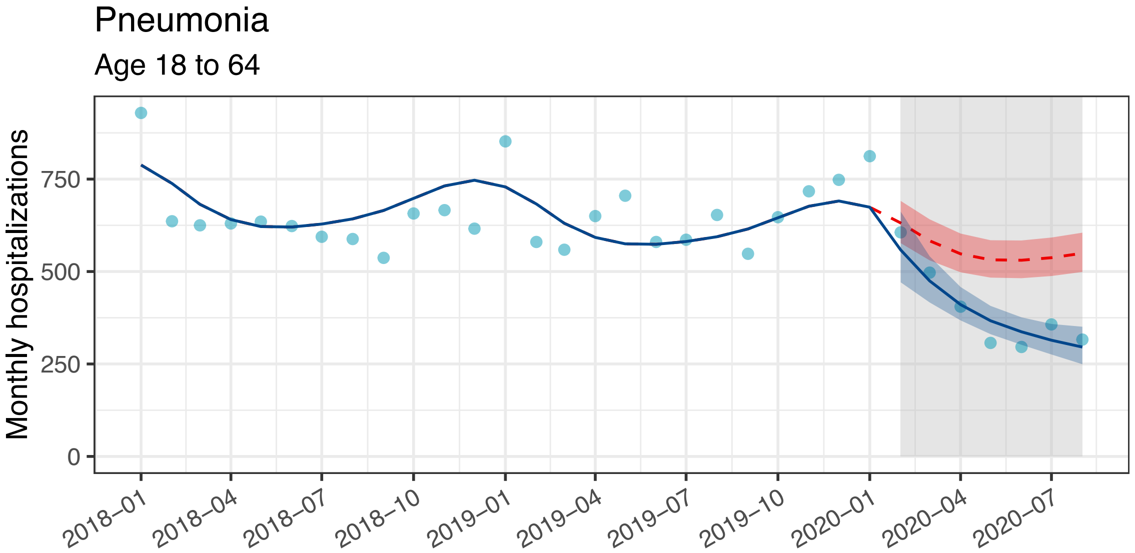
Pneumonia
Age 18 to 64
Monthly hospitalizations
0
250
500
750
2018–01
2018–04
2018–07
2018–10
2019–01
2019–04
2019–07
2019–10
2020–01
2020–04
2020–07

E

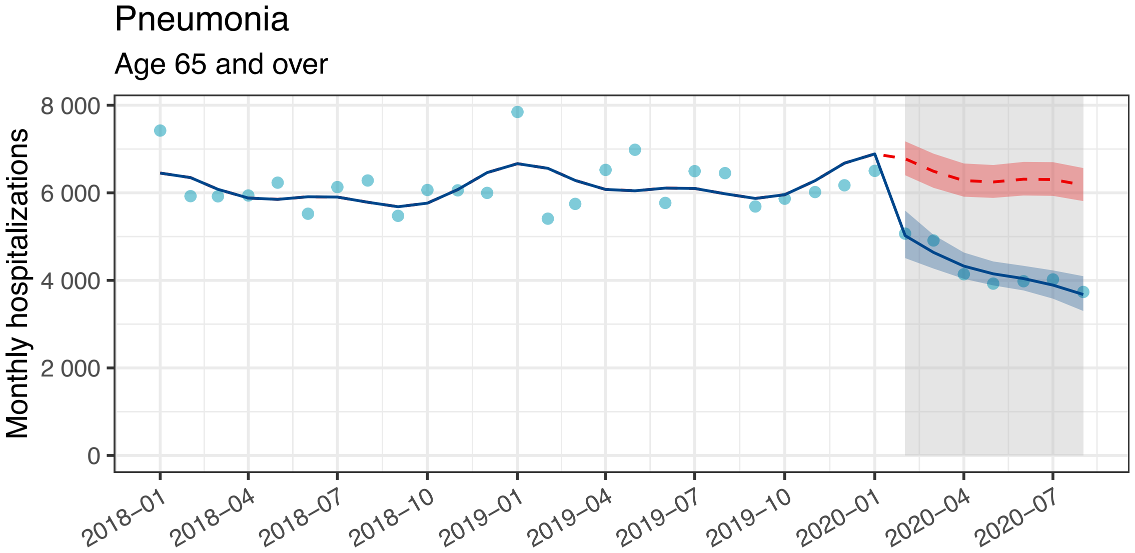
Pneumonia
Age 65 and over
Monthly hospitalizations
0
2 000
4 000
6 000
8 000
2018–01
2018–04
2018–07
2018–10
2019–01
2019–04
2019–07
2019–10
2020–01
2020–04
2020–07

F

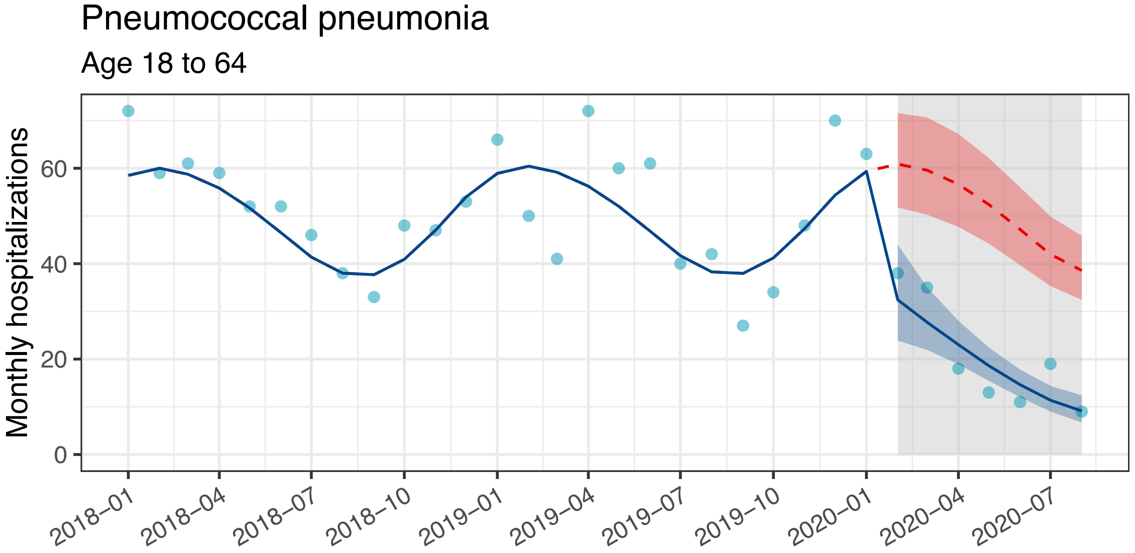
Pneumococcal pneumonia
Age 18 to 64
Monthly hospitalizations
0
20
40
60
2018–01
2018–04
2018–07
2018–10
2019–01
2019–04
2019–07
2019–10
2020–01
2020–04
2020–07

G

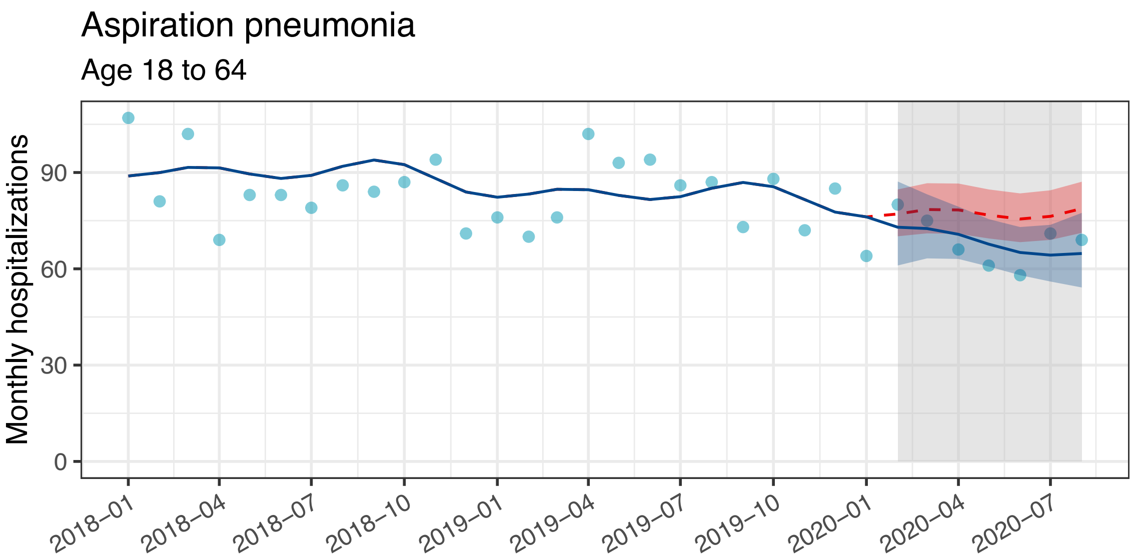
Aspiration pneumonia
Age 18 to 64
Monthly hospitalizations
0
30
60
90
2018–01
2018–04
2018–07
2018–10
2019–01
2019–04
2019–07
2019–10
2020–01
2020–04
2020–07

H

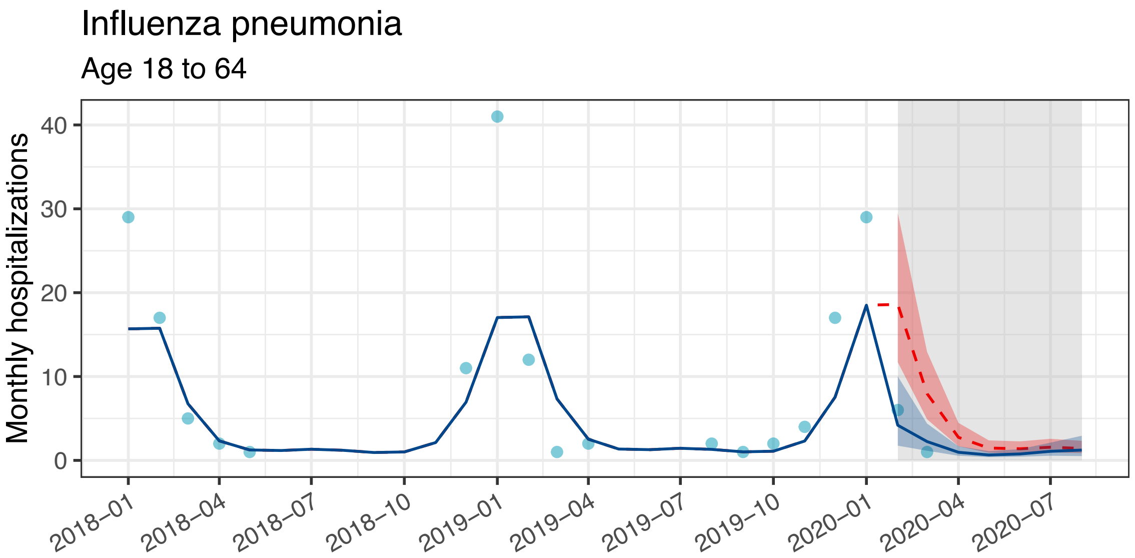
Influenza pneumonia
Age 18 to 64
Monthly hospitalizations
0
10
20
30
40
2018–01
2018–04
2018–07
2018–10
2019–01
2019–04
2019–07
2019–10
2020–01
2020–04
2020–07

I

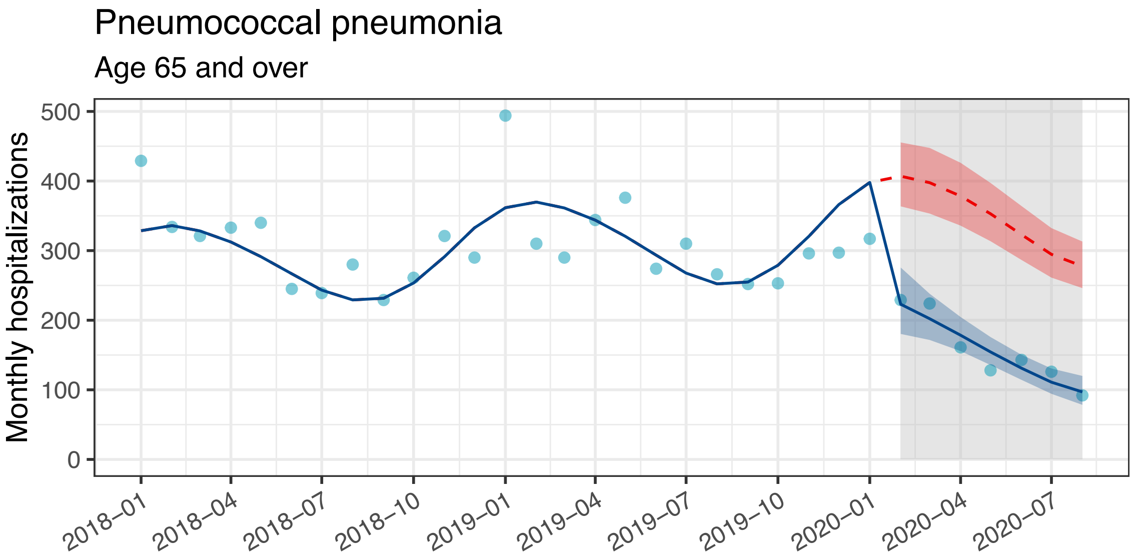
Pneumococcal pneumonia
Age 65 and over
Monthly hospitalizations
0
100
200
300
400
500
2018–01
2018–04
2018–07
2018–10
2019–01
2019–04
2019–07
2019–10
2020–01
2020–04
2020–07

J

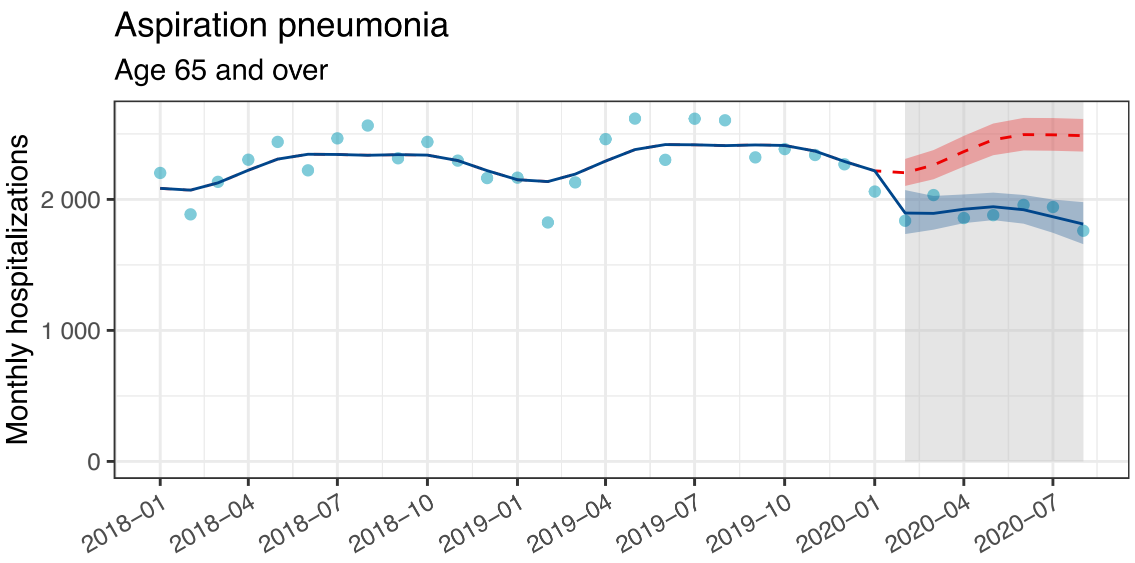
Aspiration pneumonia
Age 65 and over
Monthly hospitalizations
2 000
1 000
0
2018–01
2018–04
2018–07
2018–10
2019–01
2019–04
2019–07
2019–10
2020–01
2020–04
2020–07

K

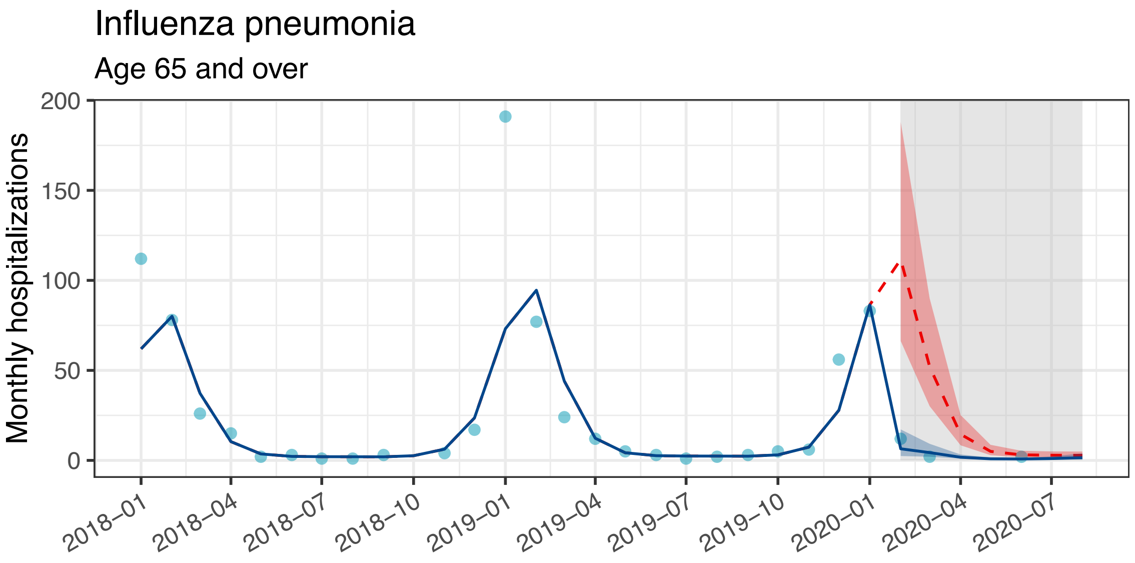
Influenza pneumonia
Age 65 and over
Monthly hospitalizations
200
150
100
50
0
2018–01
2018–04
2018–07
2018–10
2019–01
2019–04
2019–07
2019–10
2020–01
2020–04
2020–07

**Outcome: The number of 30-day deaths**

L

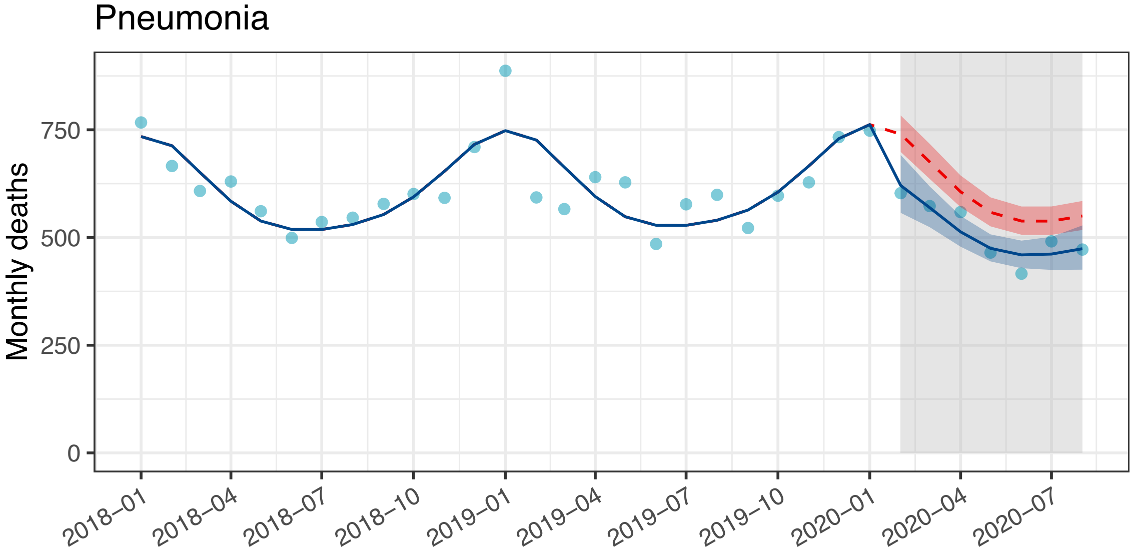


M

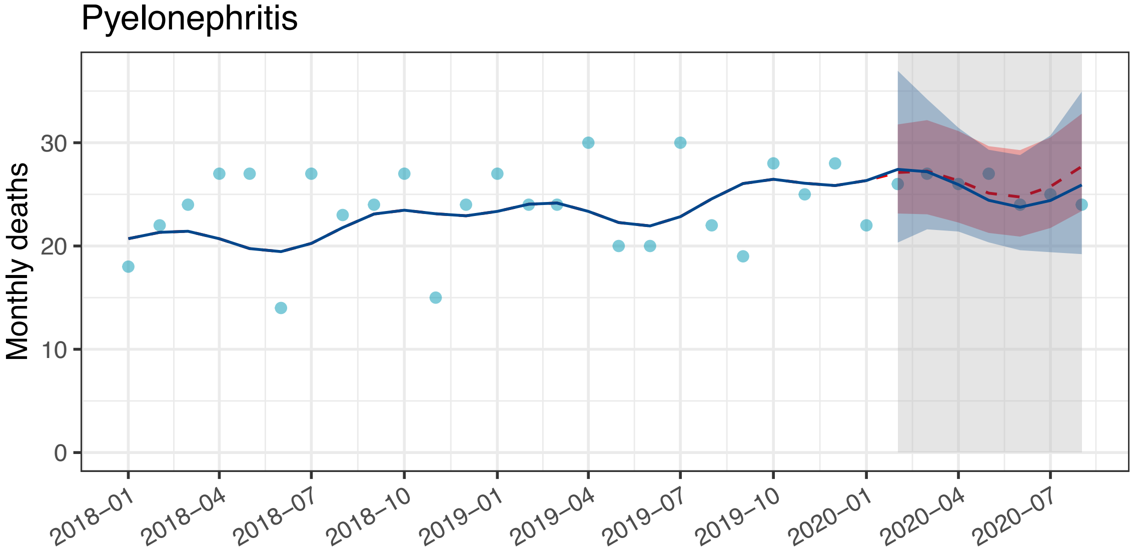


N

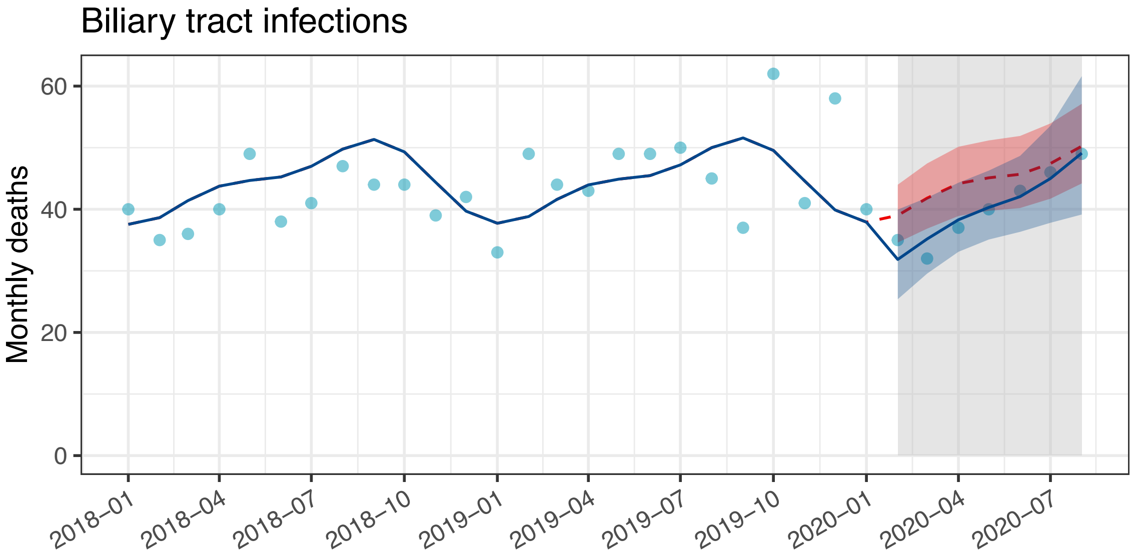

O

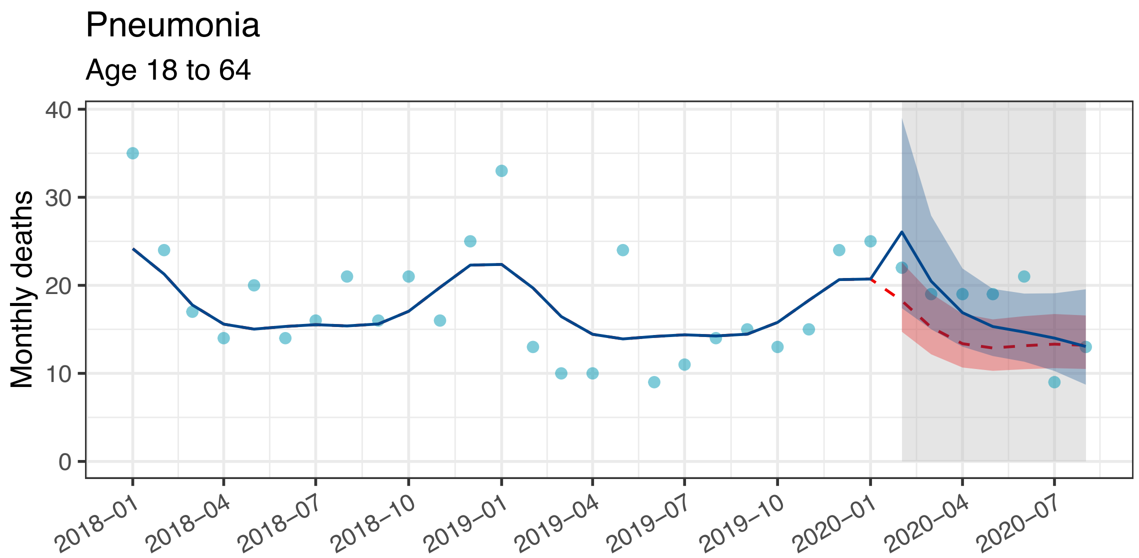

Pneumonia
Age 18 to 64
Monthly deaths
0
10
20
30
40
2018–01
2018–04
2018–07
2018–10
2019–01
2019–04
2019–07
2019–10
2020–01
2020–04
2020–07


P

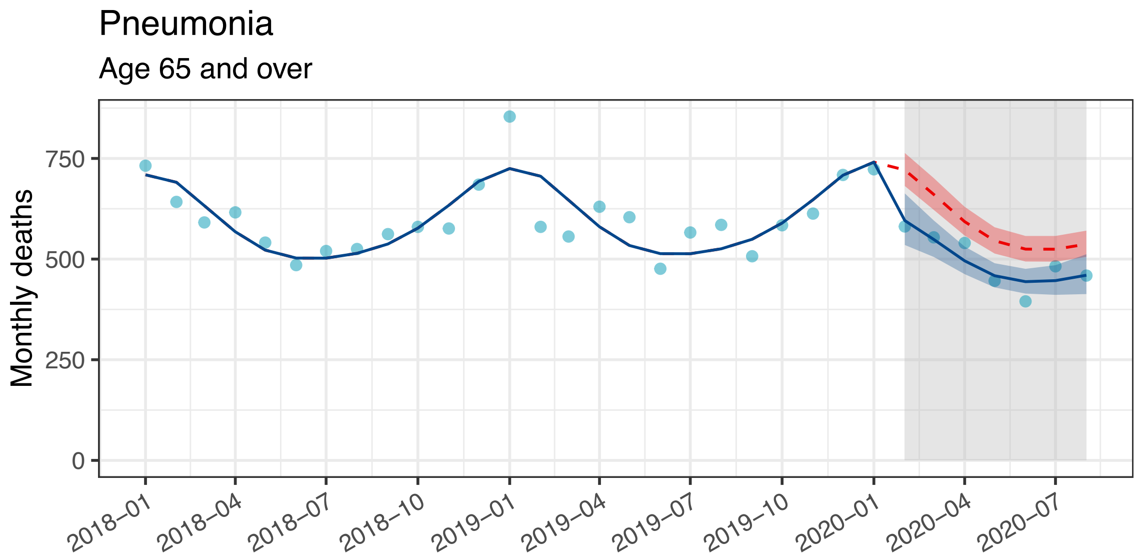

Pneumonia
Age 65 and over
Monthly deaths
0
250
500
750
2018–01
2018–04
2018–07
2018–10
2019–01
2019–04
2019–07
2019–10
2020–01
2020–04
2020–07


Q

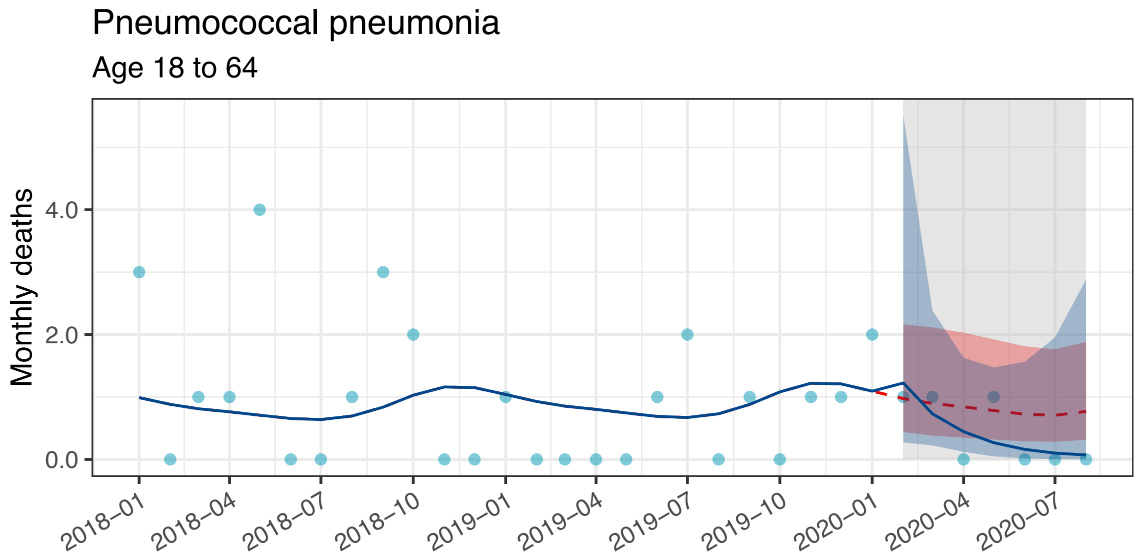

Pneumococcal pneumonia
Age 18 to 64
Monthly deaths
0.0
2.0
4.0
2018–01
2018–04
2018–07
2018–10
2019–01
2019–04
2019–07
2019–10
2020–01
2020–04
2020–07

R

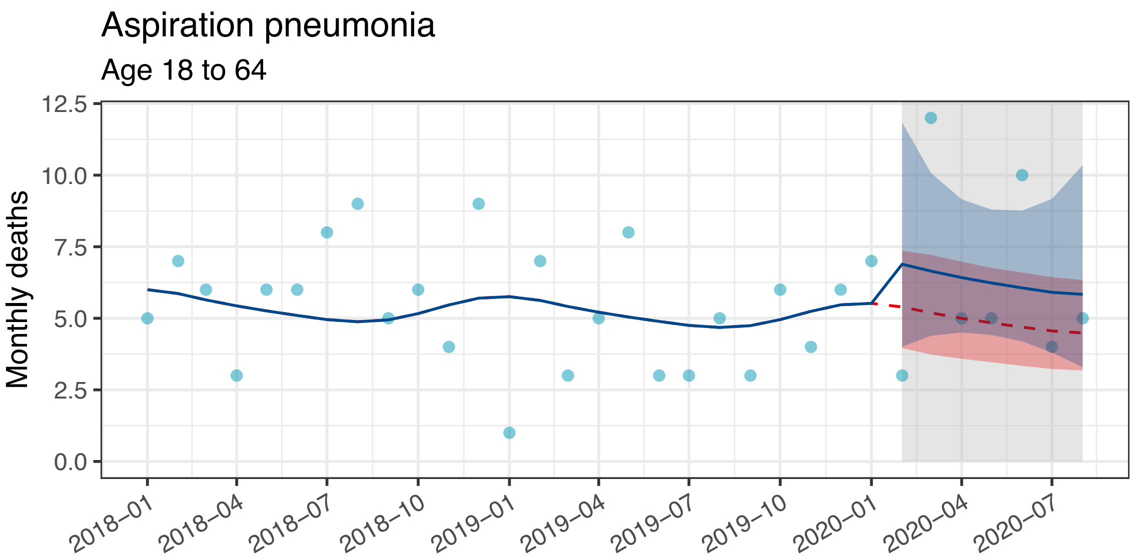
Aspiration pneumonia
Age 18 to 64
Monthly deaths
12.5
10.0
7.5
5.0
2.5
0.0
2018–01
2018–04
2018–07
2018–10
2019–01
2019–04
2019–07
2019–10
2020–01
2020–04
2020–07

S

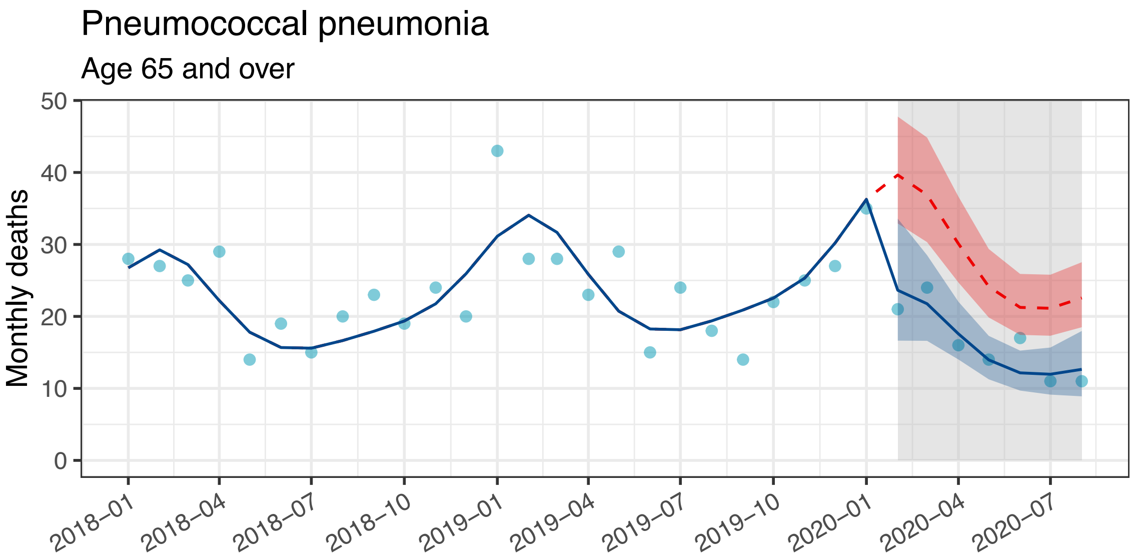
Pneumococcal pneumonia
Age 65 and over
Monthly deaths
50
40
30
20
10
0
2018–01
2018–04
2018–07
2018–10
2019–01
2019–04
2019–07
2019–10
2020–01
2020–04
2020–07

T

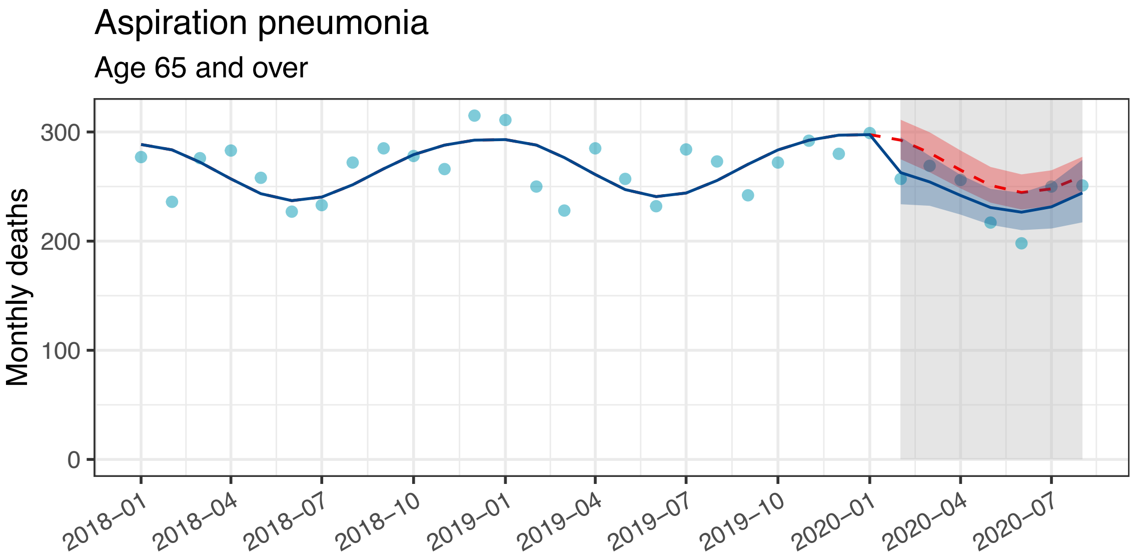
Aspiration pneumonia
Age 65 and over
Monthly deaths
300
200
100
0
2018–01
2018–04
2018–07
2018–10
2019–01
2019–04
2019–07
2019–10
2020–01
2020–04
2020–07

### SUPPLEMENTAL TABLE 2. Estimates and BIC for all models

**Outcome: The number of hospitalizations**

| Disease | Age, years | Pneumonia type | Changepoint | BIC | Δ BIC | Estimated level change rate % | (95% CI) |
|---|---|---|---|---|---|---|---|
| Pneumonia | ≧ 18 | | Null | -56·51 | 65·37 | - | - |
| | | | February | -121·88 | 0 | -24·3 | (-32·8 to -14·8) |
| | | | March | -110·98 | 10·9 | -26·5 | (-35·9 to -15·8) |
| | | | April | -105·21 | 16·67 | -32·4 | (-41·8 to -21·3) |
| Pyelonephritis | ≧ 18 | | Null | -171·27 | 1·2 | - | - |
| | | | February | -170·77 | 1·7 | -5·4 | (-12·8 to 2·6) † |
| | | | March | -168·89 | 3·58 | -3·4 | (-11·5 to 5·4) † |
| | | | April | -172·47 | 0 | -10·1 | (-17·8 to -1·6) † |
| Biliary tract infections | ≧ 18 | | Null | -164·17 | 0 | - | - |
| | | | February | -156·36 | 7·81 | -1·1 | (-9·7 to 8·3) † |
| | | | March | -156·3 | 7·87 | -0·2 | (-9·4 to 9·8) † |
| | | | April | -156·93 | 7·24 | -4·2 | (-13·5 to 6·0) † |
| Pneumonia | 18-64 | | Null | -29·78 | 33·85 | - | - |
| | | | February | -62·34 | 1·29 | -11·6 | (-26·7 to 6·7) |
| | | | March | -63·32 | 0·31 | -23·2 | (-36·9 to -6·6) |
| | | | April | -63·63 | 0 | -33·6 | (-46·1 to -18·3) |
| | ≧ 65 | | Null | -55·1 | 66·76 | - | - |
| | | | February | -121·86 | 0 | -25·9 | (-34·2 to -16·6) |
| | | | March | -108·66 | 13·2 | -27·2 | (-36·7 to -16·4) |
| | | | April | -102·29 | 19·57 | -32·7 | (-42·3 to -21·4) |
| Pneumonia | 18-64 | Pneumococcal pneumonia | Null | 72·48 | 58·99 | - | - |
| | | | February | 13·49 | 0 | -46·7 | (-62·0 to -25·5) |
| | | | March | 16·84 | 3·35 | -57·1 | (-70·2 to -38·2) |
| | | | April | 14·68 | 1·19 | -70·7 | (-79·9 to -57·1) |
| | | Aspiration pneumonia | Null | -59·51 | 0 | - | - |
| | | | February | -56·48 | 3·03 | -5·4 | (-22·2 to 15·2) |
| | | | March | -57·63 | 1·88 | -14·1 | (-30·0 to 5·5) † |
| | | | April | -59·1 | 0·41 | -21·2 | (-36·5 to -2·3) † |
| | | Influenza pneumonia | Null | 152·67 | 2·77 | - | - |
| | | | February | 149·9 | 0 | -77·5 | (-91·4 to -41·1) |

| Disease | Age, years | Pneumonia type | Changepoint | BIC | Δ BIC | Estimated level change rate % | (95% CI) |
|---|---|---|---|---|---|---|---|
| | | | March | 152·38 | 2·48 | -76·4 | (-91·6 to -33·6) |
| | | | April | 160·07 | 10·17 | -38·8 | (-80·9 to 95·6) |
| | ≧ 65 | Pneumococcal pneumonia | Null | 35·35 | 69·06 | - | - |
| | | | February | -33·71 | 0 | -45·2 | (-56·6 to -30·8) † |
| | | | March | -20·48 | 13·23 | -47·7 | (-60·2 to -31·2) † |
| | | | April | -13·25 | 20·46 | -54·9 | (-66·8 to -38·7) † |
| | | Aspiration pneumonia | Null | -108·22 | 39·5 | - | - |
| | | | February | -147·72 | 0 | -14·0 | (-21·9 to -5·2) |
| | | | March | -140·11 | 7·61 | -12·9 | (-21·9 to -2·9) † |
| | | | April | -140·15 | 7·57 | -19·3 | (-28·1 to -9·4) † |
| | | Influenza pneumonia | Null | 187·18 | 21·73 | - | - |
| | | | February | 165·45 | 0 | -94·2 | (-98·0 to -83·0) |
| | | | March | 173·28 | 7·83 | -93·7 | (-98·1 to -78·9) |
| | | | April | 188·2 | 22·75 | -82·7 | (-95·9 to -26·8) |

†: A Breusch-Godfrey test for autocorrelation was P < 0·05, indicating that the time series have autocorrelation. Confidence interval may be underestimated and thus should be interpreted with caution.

Abbreviation: BIC: Bayesian information criterion; ΔBIC: difference in BIC from the best model; CI, confidence interval

**Outcome: The number of 30-day deaths**

| Disease | Age, years | Pneumonia type | Change point | BIC | ΔBIC | Estimated level change rate % | (95% CI) |
|---|---|---|---|---|---|---|---|
| Pneumonia | ≧ 18 | | Null | -111·44 | 10·18 | - | - |
| | | | February | -121·62 | 0 | -16·1 | (-25·5 to -5·5) |
| | | | March | -115 | 6·62 | -12·3 | (-23·2 to 0·1) |
| | | | April | -112·01 | 9·61 | -10·7 | (-22·6 to 3·0) |
| Pyelonephritis | ≧ 18 | | Null | 2·49 | 0 | - | - |
| | | | February | 10·66 | 8·17 | 1·1 | (-27·2 to 40·5) |
| | | | March | 10·55 | 8·06 | 4·3 | (-26·3 to 47·6) |
| | | | April | 10·44 | 7·95 | 5·9 | (-26·7 to 52·9) |
| Biliary tract infections | ≧ 18 | | Null | -30·39 | 0 | - | - |
| | | | February | -25·31 | 5·08 | -18·4 | (-36·4 to 4·8) |
| | | | March | -25·68 | 4·71 | -21·0 | (-39·2 to 2·8) |
| | | | April | -23·13 | 7·26 | -13·0 | (-34·5 to 15·6) |
| Pneumonia | 18-64 | | Null | 44·35 | 0 | - | - |
| | | | February | 49·71 | 5·36 | 42·9 | (-8·4 to 122·8) |
| | | | March | 49·3 | 4·95 | 50·4 | (-5·8 to 140·1) |
| | | | April | 48·51 | 4·16 | 60·3 | (-2·0 to 162·3) |
| | ≧ 65 | | Null | -109·79 | 12·71 | - | - |

| Disease | Age, years | Pneumonia type | Change point | BIC | ΔBIC | Estimated level change rate % | (95% CI) |
|---|---|---|---|---|---|---|---|
| | | | February | -122·5 | 0 | -17·4 | (-26·6 to -7·1) |
| | | | March | -114·93 | 7·57 | -13·7 | (-24·4 to -1·5) |
| | | | April | -111·49 | 11·01 | -12·4 | (-24·1 to 1·2) |
| Pneumonia | 18-64 | Pneumococcal pneumonia | Null | 176·44 | 0 | - | - |
| | | | February | 182·24 | 5·8 | 25·6 | (-82·9 to 480·7) |
| | | | March | 182·1 | 5·66 | 9·1 | (-90·4 to 493·5) |
| | | | April | 182·11 | 5·67 | -44·5 | (-98·9 to 359·9) |
| | | Aspiration pneumonia | Null | 316·81 | 0 | - | - |
| | | | February | 323·65 | 6·84 | 27·7 | (-32·2 to 129·6) |
| | | | March | 319·83 | 3·02 | 101·9 | (9·3 to 258·2) |
| | | | April | 324·77 | 7·96 | 12·5 | (-46·9 to 120·3) |
| | ≧ 65 | Pneumococcal pneumonia | Null | 152·7 | 11·35 | - | - |
| | | | February | 159·08 | 0 | -40·4 | (-59·5 to -12·2) |
| | | | March | 158·29 | 5·9 | -31·8 | (-55·5 to 4·6) |
| | | | April | 154·14 | 7·7 | -34·1 | (-58·4 to 4·3) |
| | | Aspiration pneumonia | Null | 43·03 | 0 | - | - |
| | | | February | 31·68 | 3·37 | -10·2 | (-21·0 to 2·2) |
| | | | March | 37·58 | 5·67 | -6·9 | (-18·9 to 6·9) |
| | | | April | 39·38 | 5·07 | -10·1 | (-22·3 to 4·0) |

†: A Breusch-Godfrey test for autocorrelation was $P < 0·05$, indicating that the time series have autocorrelation. Confidence interval may be underestimated and thus should be interpreted with caution.

Abbreviation: BIC: Bayesian information criterion; ΔBIC: difference in BIC from the best model; CI, confidence interval
