## Appendix and ICMJE coi disclosure for "Decreased hospitalizations and deaths from community-acquired pneumonia coincided with rising public awareness of personal precautions before the governmental containment and closure policy: A nationwide observational study in Japan": ICMJE_coi_disclosure_all_220603.pdf

### ICMJE DISCLOSURE FORM

**Date: May 25, 2022**
**Your Name: Masato Tashiro**
**Manuscript Title: Decreased hospitalizations and deaths from community-acquired pneumonia coincided with rising public awareness of personal precautions before the governmental containment and closure policy: A nationwide observational study in Japan**
**Manuscript number (if known):_______________________________________________________________**

**In the interest of transparency, we ask you to disclose all relationships/activities/interests listed below that are related to the content of your manuscript. "Related" means any relation with for-profit or not-for-profit third parties whose interests may be affected by the content of the manuscript. Disclosure represents a commitment to transparency and does not necessarily indicate a bias. If you are in doubt about whether to list a relationship/activity/interest, it is preferable that you do so.**

**The following questions apply to the author's relationships/activities/interests as they relate to the current manuscript only.**

**The author's relationships/activities/interests should be defined broadly. For example, if your manuscript pertains to the epidemiology of hypertension, you should declare all relationships with manufacturers of antihypertensive medication, even if that medication is not mentioned in the manuscript.**

**In item #1 below, report all support for the work reported in this manuscript without time limit. For all other items, the time frame for disclosure is the past 36 months.**

| | | **Name all entities with whom you have this relationship or indicate none (add rows as needed)** | **Specifications/Comments (e.g., if payments were made to you or to your institution)** |
|---|---|---|---|
| | | **Time frame: Since the initial planning of the work** | |
| 1 | All support for the present manuscript (e.g., funding, provision of study materials, medical writing, article processing charges, etc.) **No time limit for this item.** | None | |
| | | **Time frame: past 36 months** | |
| 2 | Grants or contracts from any entity (if not indicated in item #1 above). | Scholarship Donation | Merck Sharp & Dohme<br>Payments were made to my institution |
| 3 | Royalties or licenses | None | |

| | | | |
|---|---|---|---|
| 4 | Consulting fees | Consulting fee | ASAHI KASEI PHARMA CORPORATION<br>Payments were made to me. |
| 5 | Payment or honoraria for lectures, presentations, speakers bureaus, manuscript writing or educational events | Honoraria for presentations | Sumitomo Dainippon Pharma Co., Ltd.<br>Payments were made to me. |
| | | Honoraria for presentations | KYORIN Holdings, Inc.<br>Payments were made to me. |
| | | Honoraria for a presentation | Astellas Pharma Inc.<br>Payments were made to me. |
| | | Honoraria for a presentation | ONO PHARMACEUTICAL CO., LTD.<br>Payments were made to me. |
| | | Honoraria for a presentation | FUJIFILM Toyama Chemical Co., Ltd.<br>Payments were made to me. |
| | | Honoraria for a presentation | CHUGAI PHARMACEUTICAL CO., LTD.<br>Payments were made to me. |
| 6 | Payment for expert testimony | None | |
| 7 | Support for attending meetings and/or travel | None | |
| 8 | Patents planned, issued or pending | None | |
| 9 | Participation on a Data Safety Monitoring Board or Advisory Board | None | |
| 10 | Leadership or fiduciary role in other board, society, committee or advocacy group, paid or unpaid | None | |
| 11 | Stock or stock options | None | |
| 12 | Receipt of equipment, materials, drugs, medical writing, gifts or other services | None | |
| 13 | Other financial or non-financial interests | None | |

**Please place an "X" next to the following statement to indicate your agreement:**

**_ X __ I certify that I have answered every question and have not altered the wording of any of the questions on this form.**

#### ICMJE DISCLOSURE FORM

**Date: May 26, 2022**
**Your Name: Shuntaro Sato**
**Manuscript Title: Decreased hospitalizations and deaths from community-acquired pneumonia coincided with rising public awareness of personal precautions before the governmental containment and closure policy: A nationwide observational study in Japan**
**Manuscript number (if known):_______________________________________________________________**

**In the interest of transparency, we ask you to disclose all relationships/activities/interests listed below that are related to the content of your manuscript. "Related" means any relation with for-profit or not-for-profit third parties whose interests may be affected by the content of the manuscript. Disclosure represents a commitment to transparency and does not necessarily indicate a bias. If you are in doubt about whether to list a relationship/activity/interest, it is preferable that you do so.**

**The following questions apply to the author's relationships/activities/interests as they relate to the current manuscript only.**

**The author's relationships/activities/interests should be defined broadly. For example, if your manuscript pertains to the epidemiology of hypertension, you should declare all relationships with manufacturers of antihypertensive medication, even if that medication is not mentioned in the manuscript.**

**In item #1 below, report all support for the work reported in this manuscript without time limit. For all other items, the time frame for disclosure is the past 36 months.**

| | | **Name all entities with whom you have this relationship or indicate none (add rows as needed)** | **Specifications/Comments (e.g., if payments were made to you or to your institution)** |
|---|---|---|---|
| | | **Time frame: Since the initial planning of the work** | |
| 1 | All support for the present manuscript (e.g., funding, provision of study materials, medical writing, article processing charges, etc.) **No time limit for this item.** | None | |
| | | **Time frame: past 36 months** | |
| 2 | Grants or contracts from any entity (if not indicated in item #1 above). | Scholarship Donation | JSPS KAKENHI<br>Payments were made to my institution |
| 3 | Royalties or licenses | None | |

| | | | |
|---|---|---|---|
| 4 | Consulting fees | Consulting fee | LIFE Research Institute.<br>Payments were made to me. |
| 5 | Payment or honoraria for lectures, presentations, speakers bureaus, manuscript writing or educational events | Honoraria for manuscript writing | Ishiyaku Pub,Inc.<br>Payments were made to me. |
| | | Honoraria for lectures | University of Tsukuba.<br>Payments were made to me. |
| | | Honoraria for lectures | Kurume university.<br>Payments were made to me. |
| | | Honoraria for lectures | Kyushu university.<br>Payments were made to me. |
| | | Honoraria for lectures | Initiative for Clinical Epidemiological Research.<br>Payments were made to me. |
| | | Honoraria for lectures | NOB DATA, Inc..<br>Payments were made to me. |
| 6 | Payment for expert testimony | None | |
| 7 | Support for attending meetings and/or travel | None | |
| 8 | Patents planned, issued or pending | None | |
| 9 | Participation on a Data Safety Monitoring Board or Advisory Board | None | |
| 10 | Leadership or fiduciary role in other board, society, committee or advocacy group, paid or unpaid | None | |
| 11 | Stock or stock options | None | |
| 12 | Receipt of equipment, materials, drugs, medical writing, gifts or other services | None | |
| 13 | Other financial or non-financial interests | None | |

**Please place an "X" next to the following statement to indicate your agreement:**

**_ X __ I certify that I have answered every question and have not altered the wording of any of the questions on this form.**

#### ICMJE DISCLOSURE FORM

**Date: May 30, 2022**
**Your Name: Akira Endo**
**Manuscript Title: Decreased hospitalizations and deaths from community-acquired pneumonia coincided with rising public awareness of personal precautions before the governmental containment and closure policy: A nationwide observational study in Japan**
**Manuscript number (if known):_______________________________________________________________**

**In the interest of transparency, we ask you to disclose all relationships/activities/interests listed below that are related to the content of your manuscript. "Related" means any relation with for-profit or not-for-profit third parties whose interests may be affected by the content of the manuscript. Disclosure represents a commitment to transparency and does not necessarily indicate a bias. If you are in doubt about whether to list a relationship/activity/interest, it is preferable that you do so.**

**The following questions apply to the author's relationships/activities/interests as they relate to the current manuscript only.**

**The author's relationships/activities/interests should be defined broadly. For example, if your manuscript pertains to the epidemiology of hypertension, you should declare all relationships with manufacturers of antihypertensive medication, even if that medication is not mentioned in the manuscript.**

**In item #1 below, report all support for the work reported in this manuscript without time limit. For all other items, the time frame for disclosure is the past 36 months.**

| | | **Name all entities with whom you have this relationship or indicate none (add rows as needed)** | **Specifications/Comments (e.g., if payments were made to you or to your institution)** |
|---|---|---|---|
| **Time frame: Since the initial planning of the work** | | | |
| 1 | All support for the present manuscript (e.g., funding, provision of study materials, medical writing, article processing charges, etc.) **No time limit for this item.** | JSPS Grants-in-aid (KAKENHI) | Payments were made to my institution |
| **Time frame: past 36 months** | | | |
| 2 | Grants or contracts from any entity (if not indicated in item #1 above). | Taisho Pharmaceutical Co., Ltd. | Payments were made to me |
| 3 | Royalties or licenses | None | |

| | | |
|---|---|---|
| 4 | Consulting fees | None |
| 5 | Payment or honoraria for lectures, presentations, speakers bureaus, manuscript writing or educational events | None |
| 6 | Payment for expert testimony | None |
| 7 | Support for attending meetings and/or travel | None |
| 8 | Patents planned, issued or pending | None |
| 9 | Participation on a Data Safety Monitoring Board or Advisory Board | None |
| 10 | Leadership or fiduciary role in other board, society, committee or advocacy group, paid or unpaid | None |
| 11 | Stock or stock options | None |
| 12 | Receipt of equipment, materials, drugs, medical writing, gifts or other services | None |
| 13 | Other financial or non-financial interests | None |

**Please place an "X" next to the following statement to indicate your agreement:**

**_ X __ I certify that I have answered every question and have not altered the wording of any of the questions on this form.**

### ICMJE DISCLOSURE FORM

**Date: May 27, 2022**
**Your Name: Ryosuke Hamashima**
**Manuscript Title: Decreased hospitalizations and deaths from community-acquired pneumonia coincided with rising public awareness of personal precautions before the governmental containment and closure policy: A nationwide observational study in Japan**
**Manuscript number (if known):_______________________________________________________________**

**In the interest of transparency, we ask you to disclose all relationships/activities/interests listed below that are related to the content of your manuscript. "Related" means any relation with for-profit or not-for-profit third parties whose interests may be affected by the content of the manuscript. Disclosure represents a commitment to transparency and does not necessarily indicate a bias. If you are in doubt about whether to list a relationship/activity/interest, it is preferable that you do so.**

**The following questions apply to the author's relationships/activities/interests as they relate to the current manuscript only.**

**The author's relationships/activities/interests should be defined broadly. For example, if your manuscript pertains to the epidemiology of hypertension, you should declare all relationships with manufacturers of antihypertensive medication, even if that medication is not mentioned in the manuscript.**

**In item #1 below, report all support for the work reported in this manuscript without time limit. For all other items, the time frame for disclosure is the past 36 months.**

| | | **Name all entities with whom you have this relationship or indicate none (add rows as needed)** | **Specifications/Comments (e.g., if payments were made to you or to your institution)** |
|---|---|---|---|
| | | **Time frame: Since the initial planning of the work** | |
| 1 | All support for the present manuscript (e.g., funding, provision of study materials, medical writing, article processing charges, etc.) **No time limit for this item.** | None | |
| | | **Time frame: past 36 months** | |
| 2 | Grants or contracts from any entity (if not indicated in item #1 above). | None | |
| 3 | Royalties or licenses | None | |

| | | | |
|---|---|---|---|
| 4 | Consulting fees | None | |
| 5 | Payment or honoraria for lectures, presentations, speakers bureaus, manuscript writing or educational events | Honoraria for presentations | AstraZeneca plc.<br>Payments were made to me. |
| 6 | Payment for expert testimony | None | |
| 7 | Support for attending meetings and/or travel | None | |
| 8 | Patents planned, issued or pending | None | |
| 9 | Participation on a Data Safety Monitoring Board or Advisory Board | None | |
| 10 | Leadership or fiduciary role in other board, society, committee or advocacy group, paid or unpaid | None | |
| 11 | Stock or stock options | None | |
| 12 | Receipt of equipment, materials, drugs, medical writing, gifts or other services | None | |
| 13 | Other financial or non-financial interests | None | |

**Please place an "X" next to the following statement to indicate your agreement:**

**_ X __ I certify that I have answered every question and have not altered the wording of any of the questions on this form.**

**ICMJE DISCLOSURE FORM**

**Date: May 26, 2022**
**Your Name: Yuya Ito**
**Manuscript Title: Decreased hospitalizations and deaths from community-acquired pneumonia coincided with rising public awareness of personal precautions before the governmental containment and closure policy: A nationwide observational study in Japan**
**Manuscript number (if known):_________________________________________________________________________**

**In the interest of transparency, we ask you to disclose all relationships/activities/interests listed below that are related to the content of your manuscript. "Related" means any relation with for-profit or not-for-profit third parties whose interests may be affected by the content of the manuscript. Disclosure represents a commitment to transparency and does not necessarily indicate a bias. If you are in doubt about whether to list a relationship/activity/interest, it is preferable that you do so.**

**The following questions apply to the author's relationships/activities/interests as they relate to the current manuscript only.**

**The author's relationships/activities/interests should be defined broadly. For example, if your manuscript pertains to the epidemiology of hypertension, you should declare all relationships with manufacturers of antihypertensive medication, even if that medication is not mentioned in the manuscript.**

**In item #1 below, report all support for the work reported in this manuscript without time limit. For all other items, the time frame for disclosure is the past 36 months.**

| | | **Name all entities with whom you have this relationship or indicate none (add rows as needed)** | **Specifications/Comments (e.g., if payments were made to you or to your institution)** |
|---|---|---|---|
| **Time frame: Since the initial planning of the work** | | | |
| 1 | All support for the present manuscript (e.g., funding, provision of study materials, medical writing, article processing charges, etc.) **No time limit for this item.** | None | |
| **Time frame: past 36 months** | | | |
| 2 | Grants or contracts from any entity (if not indicated in item #1 above). | None | |
| 3 | Royalties or licenses | None | |
| 4 | Consulting fees | None | |

| | | | |
|---|---|---|---|
| 5 | Payment or honoraria for lectures, presentations, speakers bureaus, manuscript writing or educational events | Honoraria for lecture. | CHUGAI PHARMACEUTICAL CO., LTD.<br>Payments were made to me. |
| | | Honoraria for lecture and presentation. | AstraZeneca Pharmaceuticals Co., Ltd.<br>Payments were made to me. |
| | | Honoraria for manuscript writing. | Japan medical journal.<br>Payments were made to me. |
| 6 | Payment for expert testimony | None | |
| 7 | Support for attending meetings and/or travel | None | |
| 8 | Patents planned, issued or pending | None | |
| 9 | Participation on a Data Safety Monitoring Board or Advisory Board | None | |
| 10 | Leadership or fiduciary role in other board, society, committee or advocacy group, paid or unpaid | None | |
| 11 | Stock or stock options | None | |
| 12 | Receipt of equipment, materials, drugs, medical writing, gifts or other services | None | |
| 13 | Other financial or non-financial interests | None | |

**Please place an "X" next to the following statement to indicate your agreement:**

**_ X __ I certify that I have answered every question and have not altered the wording of any of the questions on this form.**

#### ICMJE DISCLOSURE FORM

**Date: May 26, 2022**
**Your Name: Nobuyuki Ashizawa**
**Manuscript Title: Decreased hospitalizations and deaths from community-acquired pneumonia coincided with rising public awareness of personal precautions before the governmental containment and closure policy: A nationwide observational study in Japan**
**Manuscript number (if known):______________________________________________________________**

**In the interest of transparency, we ask you to disclose all relationships/activities/interests listed below that are related to the content of your manuscript. "Related" means any relation with for-profit or not-for-profit third parties whose interests may be affected by the content of the manuscript. Disclosure represents a commitment to transparency and does not necessarily indicate a bias. If you are in doubt about whether to list a relationship/activity/interest, it is preferable that you do so.**

**The following questions apply to the author's relationships/activities/interests as they relate to the current manuscript only.**

**The author's relationships/activities/interests should be defined broadly. For example, if your manuscript pertains to the epidemiology of hypertension, you should declare all relationships with manufacturers of antihypertensive medication, even if that medication is not mentioned in the manuscript.**

**In item #1 below, report all support for the work reported in this manuscript without time limit. For all other items, the time frame for disclosure is the past 36 months.**

| | | **Name all entities with whom you have this relationship or indicate none (add rows as needed)** | **Specifications/Comments (e.g., if payments were made to you or to your institution)** |
|---|---|---|---|
| | | **Time frame: Since the initial planning of the work** | |
| 1 | All support for the present manuscript (e.g., funding, provision of study materials, medical writing, article processing charges, etc.) **No time limit for this item.** | None | |
| | | **Time frame: past 36 months** | |
| 2 | Grants or contracts from any entity (if not indicated in item #1 above). | None | |
| 3 | Royalties or licenses | None | |
| 4 | Consulting fees | None | |

| | | | |
|---|---|---|---|
| 5 | Payment or honoraria for lectures, presentations, speakers bureaus, manuscript writing or educational events | Honoraria for presentations | ASAHI KASEI PHARMA CORPORATION<br>Payments were made to me. |
| 6 | Payment for expert testimony | None | |
| 7 | Support for attending meetings and/or travel | None | |
| 8 | Patents planned, issued or pending | None | |
| 9 | Participation on a Data Safety Monitoring Board or Advisory Board | None | |
| 10 | Leadership or fiduciary role in other board, society, committee or advocacy group, paid or unpaid | None | |
| 11 | Stock or stock options | None | |
| 12 | Receipt of equipment, materials, drugs, medical writing, gifts or other services | None | |
| 13 | Other financial or non-financial interests | None | |

**Please place an "X" next to the following statement to indicate your agreement:**

**_ X __ I certify that I have answered every question and have not altered the wording of any of the questions on this form.**

#### ICMJE DISCLOSURE FORM

**Date: May 30, 2022**
**Your Name: Kazuaki Takeda**
**Manuscript Title: Decreased hospitalizations and deaths from community-acquired pneumonia coincided with rising public awareness of personal precautions before the governmental containment and closure policy: A nationwide observational study in Japan**
**Manuscript number (if known):_______________________________________________________________**

**In the interest of transparency, we ask you to disclose all relationships/activities/interests listed below that are related to the content of your manuscript. "Related" means any relation with for-profit or not-for-profit third parties whose interests may be affected by the content of the manuscript. Disclosure represents a commitment to transparency and does not necessarily indicate a bias. If you are in doubt about whether to list a relationship/activity/interest, it is preferable that you do so.**

**The following questions apply to the author's relationships/activities/interests as they relate to the current manuscript only.**

**The author's relationships/activities/interests should be defined broadly. For example, if your manuscript pertains to the epidemiology of hypertension, you should declare all relationships with manufacturers of antihypertensive medication, even if that medication is not mentioned in the manuscript.**

**In item #1 below, report all support for the work reported in this manuscript without time limit. For all other items, the time frame for disclosure is the past 36 months.**

| | | **Name all entities with whom you have this relationship or indicate none (add rows as needed)** | **Specifications/Comments (e.g., if payments were made to you or to your institution)** |
|---|---|---|---|
| | | **Time frame: Since the initial planning of the work** | |
| 1 | All support for the present manuscript (e.g., funding, provision of study materials, medical writing, article processing charges, etc.) **No time limit for this item.** | None | |
| | | **Time frame: past 36 months** | |
| 2 | Grants or contracts from any entity (if not indicated in item #1 above). | None | |
| 3 | Royalties or licenses | None | |
| 4 | Consulting fees | None | |

| | | |
|---|---|---|
| 5 | Payment or honoraria for lectures, presentations, speakers bureaus, manuscript writing or educational events | None |
| 6 | Payment for expert testimony | None |
| 7 | Support for attending meetings and/or travel | None |
| 8 | Patents planned, issued or pending | None |
| 9 | Participation on a Data Safety Monitoring Board or Advisory Board | None |
| 10 | Leadership or fiduciary role in other board, society, committee or advocacy group, paid or unpaid | None |
| 11 | Stock or stock options | None |
| 12 | Receipt of equipment, materials, drugs, medical writing, gifts or other services | None |
| 13 | Other financial or non-financial interests | None |

**Please place an "X" next to the following statement to indicate your agreement:**

**_ X __ I certify that I have answered every question and have not altered the wording of any of the questions on this form.**

#### ICMJE DISCLOSURE FORM

**Date: May 26, 2022**
**Your Name: Naoki Iwanaga**
**Manuscript Title: Decreased hospitalizations and deaths from community-acquired pneumonia coincided with rising public awareness of personal precautions before the governmental containment and closure policy: A nationwide observational study in Japan**
**Manuscript number (if known):________________________________________________________________**

**In the interest of transparency, we ask you to disclose all relationships/activities/interests listed below that are related to the content of your manuscript. "Related" means any relation with for-profit or not-for-profit third parties whose interests may be affected by the content of the manuscript. Disclosure represents a commitment to transparency and does not necessarily indicate a bias. If you are in doubt about whether to list a relationship/activity/interest, it is preferable that you do so.**

**The following questions apply to the author's relationships/activities/interests as they relate to the current manuscript only.**

**The author's relationships/activities/interests should be defined broadly. For example, if your manuscript pertains to the epidemiology of hypertension, you should declare all relationships with manufacturers of antihypertensive medication, even if that medication is not mentioned in the manuscript.**

**In item #1 below, report all support for the work reported in this manuscript without time limit. For all other items, the time frame for disclosure is the past 36 months.**

| | | **Name all entities with whom you have this relationship or indicate none (add rows as needed)** | **Specifications/Comments (e.g., if payments were made to you or to your institution)** |
|---|---|---|---|
| | | **Time frame: Since the initial planning of the work** | |
| 1 | All support for the present manuscript (e.g., funding, provision of study materials, medical writing, article processing charges, etc.) **No time limit for this item.** | None | |
| | | **Time frame: past 36 months** | |
| 2 | Grants or contracts from any entity (if not indicated in item #1 above). | None | |
| 3 | Royalties or licenses | None | |
| 4 | Consulting fees | None | |

| | | | |
|---|---|---|---|
| 5 | Payment or honoraria for lectures, presentations, speakers bureaus, manuscript writing or educational events | Honoraria for presentations | Merck Sharp & Dohme<br>Payments were made to me. |
| | | Honoraria for presentations | TAISHO PHARMACEUTICAL CO., LTD.<br>Payments were made to me. |
| 6 | Payment for expert testimony | None | |
| 7 | Support for attending meetings and/or travel | None | |
| 8 | Patents planned, issued or pending | None | |
| 9 | Participation on a Data Safety Monitoring Board or Advisory Board | None | |
| 10 | Leadership or fiduciary role in other board, society, committee or advocacy group, paid or unpaid | None | |
| 11 | Stock or stock options | None | |
| 12 | Receipt of equipment, materials, drugs, medical writing, gifts or other services | None | |
| 13 | Other financial or non-financial interests | None | |

**Please place an "X" next to the following statement to indicate your agreement:**

**_ X __ I certify that I have answered every question and have not altered the wording of any of the questions on this form.**

### ICMJE DISCLOSURE FORM

**Date: May 26, 2022**
**Your Name: Shotaro Ide**
**Manuscript Title: Decreased hospitalizations and deaths from community-acquired pneumonia coincided with rising public awareness of personal precautions before the governmental containment and closure policy: A nationwide observational study in Japan**
**Manuscript number (if known):______________________________________________________________**

**In the interest of transparency, we ask you to disclose all relationships/activities/interests listed below that are related to the content of your manuscript. "Related" means any relation with for-profit or not-for-profit third parties whose interests may be affected by the content of the manuscript. Disclosure represents a commitment to transparency and does not necessarily indicate a bias. If you are in doubt about whether to list a relationship/activity/interest, it is preferable that you do so.**

**The following questions apply to the author's relationships/activities/interests as they relate to the current manuscript only.**

**The author's relationships/activities/interests should be defined broadly. For example, if your manuscript pertains to the epidemiology of hypertension, you should declare all relationships with manufacturers of antihypertensive medication, even if that medication is not mentioned in the manuscript.**

**In item #1 below, report all support for the work reported in this manuscript without time limit. For all other items, the time frame for disclosure is the past 36 months.**

| | | **Name all entities with whom you have this relationship or indicate none (add rows as needed)** | **Specifications/Comments (e.g., if payments were made to you or to your institution)** |
|---|---|---|---|
| | | **Time frame: Since the initial planning of the work** | |
| 1 | All support for the present manuscript (e.g., funding, provision of study materials, medical writing, article processing charges, etc.) **No time limit for this item.** | None | |
| | | **Time frame: past 36 months** | |
| 2 | Grants or contracts from any entity (if not indicated in item #1 above). | None | |
| 3 | Royalties or licenses | None | |
| 4 | Consulting fees | None | |

| | | | |
|---|---|---|---|
| 5 | Payment or honoraria for lectures, presentations, speakers bureaus, manuscript writing or educational events | Honoraria for presentations | KYORIN Holdings, Inc.<br>Payments were made to me. |
| | | Honoraria for presentations | AstraZeneca Holdings, Inc.<br>Payments were made to me. |
| 6 | Payment for expert testimony | None | |
| 7 | Support for attending meetings and/or travel | None | |
| 8 | Patents planned, issued or pending | None | |
| 9 | Participation on a Data Safety Monitoring Board or Advisory Board | None | |
| 10 | Leadership or fiduciary role in other board, society, committee or advocacy group, paid or unpaid | None | |
| 11 | Stock or stock options | None | |
| 12 | Receipt of equipment, materials, drugs, medical writing, gifts or other services | None | |
| 13 | Other financial or non-financial interests | None | |

**Please place an "X" next to the following statement to indicate your agreement:**

**_ X _ I certify that I have answered every question and have not altered the wording of any of the questions on this form.**

#### ICMJE DISCLOSURE FORM

**Date: May 28, 2022**
**Your Name: Ayumi Fujita**
**Manuscript Title: Decreased hospitalizations and deaths from community-acquired pneumonia coincided with rising public awareness of personal precautions before the governmental containment and closure policy: A nationwide observational study in Japan**
**Manuscript number (if known):________________________________________________________________**

**In the interest of transparency, we ask you to disclose all relationships/activities/interests listed below that are related to the content of your manuscript. "Related" means any relation with for-profit or not-for-profit third parties whose interests may be affected by the content of the manuscript. Disclosure represents a commitment to transparency and does not necessarily indicate a bias. If you are in doubt about whether to list a relationship/activity/interest, it is preferable that you do so.**

**The following questions apply to the author's relationships/activities/interests as they relate to the current manuscript only.**

**The author's relationships/activities/interests should be defined broadly. For example, if your manuscript pertains to the epidemiology of hypertension, you should declare all relationships with manufacturers of antihypertensive medication, even if that medication is not mentioned in the manuscript.**

**In item #1 below, report all support for the work reported in this manuscript without time limit. For all other items, the time frame for disclosure is the past 36 months.**

| | | **Name all entities with whom you have this relationship or indicate none (add rows as needed)** | **Specifications/Comments (e.g., if payments were made to you or to your institution)** |
|---|---|---|---|
| | | **Time frame: Since the initial planning of the work** | |
| 1 | All support for the present manuscript (e.g., funding, provision of study materials, medical writing, article processing charges, etc.) **No time limit for this item.** | None | |
| | | **Time frame: past 36 months** | |
| 2 | Grants or contracts from any entity (if not indicated in item #1 above). | None | |
| 3 | Royalties or licenses | None | |
| 4 | Consulting fees | None | |

| | | |
|---|---|---|
| 5 | Payment or honoraria for lectures, presentations, speakers bureaus, manuscript writing or educational events | None |
| 6 | Payment for expert testimony | None |
| 7 | Support for attending meetings and/or travel | None |
| 8 | Patents planned, issued or pending | None |
| 9 | Participation on a Data Safety Monitoring Board or Advisory Board | None |
| 10 | Leadership or fiduciary role in other board, society, committee or advocacy group, paid or unpaid | None |
| 11 | Stock or stock options | None |
| 12 | Receipt of equipment, materials, drugs, medical writing, gifts or other services | None |
| 13 | Other financial or non-financial interests | None |

**Please place an "X" next to the following statement to indicate your agreement:**

**_ X __ I certify that I have answered every question and have not altered the wording of any of the questions on this form.**

### ICMJE DISCLOSURE FORM

**Date: May 26, 2022**
**Your Name: Takahiro Takazono**
**Manuscript Title: Decreased hospitalizations and deaths from community-acquired pneumonia coincided with rising public awareness of personal precautions before the governmental containment and closure policy: A nationwide observational study in Japan**
**Manuscript number (if known):_______________________________________________________________**

**In the interest of transparency, we ask you to disclose all relationships/activities/interests listed below that are related to the content of your manuscript. "Related" means any relation with for-profit or not-for-profit third parties whose interests may be affected by the content of the manuscript. Disclosure represents a commitment to transparency and does not necessarily indicate a bias. If you are in doubt about whether to list a relationship/activity/interest, it is preferable that you do so.**

**The following questions apply to the author's relationships/activities/interests as they relate to the current manuscript only.**

**The author's relationships/activities/interests should be defined broadly. For example, if your manuscript pertains to the epidemiology of hypertension, you should declare all relationships with manufacturers of antihypertensive medication, even if that medication is not mentioned in the manuscript.**

**In item #1 below, report all support for the work reported in this manuscript without time limit. For all other items, the time frame for disclosure is the past 36 months.**

| | | **Name all entities with whom you have this relationship or indicate none (add rows as needed)** | **Specifications/Comments (e.g., if payments were made to you or to your institution)** |
|---|---|---|---|
| | | **Time frame: Since the initial planning of the work** | |
| 1 | All support for the present manuscript (e.g., funding, provision of study materials, medical writing, article processing charges, etc.) **No time limit for this item.** | None | |
| | | **Time frame: past 36 months** | |
| 2 | Grants or contracts from any entity (if not indicated in item #1 above). | None | |
| 3 | Royalties or licenses | None | |
| 4 | Consulting fees | None | |

| | | |
|---|---|---|
| 5 | Payment or honoraria for lectures, presentations, speakers bureaus, manuscript writing or educational events | None |
| 6 | Payment for expert testimony | None |
| 7 | Support for attending meetings and/or travel | None |
| 8 | Patents planned, issued or pending | None |
| 9 | Participation on a Data Safety Monitoring Board or Advisory Board | None |
| 10 | Leadership or fiduciary role in other board, society, committee or advocacy group, paid or unpaid | None |
| 11 | Stock or stock options | None |
| 12 | Receipt of equipment, materials, drugs, medical writing, gifts or other services | None |
| 13 | Other financial or non-financial interests | None |

**Please place an "X" next to the following statement to indicate your agreement:**

**_ X __ I certify that I have answered every question and have not altered the wording of any of the questions on this form.**

**ICMJE DISCLOSURE FORM**

**Date: May 30, 2022**
**Your Name: Kazuko Yamamoto**
**Manuscript Title: Decreased hospitalizations and deaths from community-acquired pneumonia coincided with rising public awareness of personal precautions before the governmental containment and closure policy: A nationwide observational study in Japan**
**Manuscript number (if known):_________________________________________________________________**

**In the interest of transparency, we ask you to disclose all relationships/activities/interests listed below that are related to the content of your manuscript. "Related" means any relation with for-profit or not-for-profit third parties whose interests may be affected by the content of the manuscript. Disclosure represents a commitment to transparency and does not necessarily indicate a bias. If you are in doubt about whether to list a relationship/activity/interest, it is preferable that you do so.**

**The following questions apply to the author's relationships/activities/interests as they relate to the current manuscript only.**

**The author's relationships/activities/interests should be defined broadly. For example, if your manuscript pertains to the epidemiology of hypertension, you should declare all relationships with manufacturers of antihypertensive medication, even if that medication is not mentioned in the manuscript.**

**In item #1 below, report all support for the work reported in this manuscript without time limit. For all other items, the time frame for disclosure is the past 36 months.**

| | | **Name all entities with whom you have this relationship or indicate none (add rows as needed)** | **Specifications/Comments (e.g., if payments were made to you or to your institution)** |
|---|---|---|---|
| | | **Time frame: Since the initial planning of the work** | |
| 1 | All support for the present manuscript (e.g., funding, provision of study materials, medical writing, article processing charges, etc.) **No time limit for this item.** | None | |
| | | **Time frame: past 36 months** | |
| 2 | Grants or contracts from any entity (if not indicated in item #1 above). | Contracts for Clinical Study | Kirin Holdings Company<br>Payments were made to my institution |
| | | Contracts for Clinical Study | Fisher & Paykel Healthcare<br>Payments were made to my institution |
| 3 | Royalties or licenses | None | |
| 4 | Consulting fees | None | |
| 5 | | Honoraria for a lecture | Astellas Pharma Inc. |

| | | | |
|---|---|---|---|
| | Payment or honoraria for lectures, presentations, speakers bureaus, manuscript writing or educational events | | Payments were made to me. |
| | | Honoraria for a lecture | Merck & Co.<br>Payments were made to me. |
| | | Honoraria for a lecture | Fisher & Paykel Healthcare<br>Payments were made to me. |
| | | Honoraria for a lecture | Senju Pharmaceutical Co., Ltd.<br>Payments were made to me. |
| | | Honoraria for a lecture | Taisho Pharmaceutical.<br>Payments were made to me. |
| | | Honoraria for a lecture | Otsuka Pharmaceutical Factory, Inc.<br>Payments were made to me. |
| | | Honoraria for a lecture | Meiji Seika Pharma. Co., Ltd.<br>Payments were made to me. |
| | | Honoraria for a lecture | Kyowa Kirin<br>Payments were made to me. |
| | | Honoraria for a lecture | FUJIFILM Toyama Chemical Co., Ltd.<br>Payments were made to me. |
| 6 | Payment for expert testimony | None | |
| 7 | Support for attending meetings and/or travel | None | |
| 8 | Patents planned, issued or pending | Patents issued and continuation | Teijin Pharma Ltd. |
| 9 | Participation on a Data Safety Monitoring Board or Advisory Board | None | |
| 10 | Leadership or fiduciary role in other board, society, committee or advocacy group, paid or unpaid | None | |
| 11 | Stock or stock options | None | |
| 12 | Receipt of equipment, materials, drugs, medical writing, gifts or other services | None | |
| 13 | Other financial or non-financial interests | None | |

**Please place an "X" next to the following statement to indicate your agreement:**

**_ X __ I certify that I have answered every question and have not altered the wording of any of the questions on this form.**

**ICMJE DISCLOSURE FORM**

**Date: May 26, 2022**
**Your Name: Takeshi Tanaka**
**Manuscript Title: Decreased hospitalizations and deaths from community-acquired pneumonia coincided with rising public awareness of personal precautions before the governmental containment and closure policy: A nationwide observational study in Japan**
**Manuscript number (if known):_________________________________________________________________**

**In the interest of transparency, we ask you to disclose all relationships/activities/interests listed below that are related to the content of your manuscript. "Related" means any relation with for-profit or not-for-profit third parties whose interests may be affected by the content of the manuscript. Disclosure represents a commitment to transparency and does not necessarily indicate a bias. If you are in doubt about whether to list a relationship/activity/interest, it is preferable that you do so.**

**The following questions apply to the author's relationships/activities/interests as they relate to the current manuscript only.**

**The author's relationships/activities/interests should be defined broadly. For example, if your manuscript pertains to the epidemiology of hypertension, you should declare all relationships with manufacturers of antihypertensive medication, even if that medication is not mentioned in the manuscript.**

**In item #1 below, report all support for the work reported in this manuscript without time limit. For all other items, the time frame for disclosure is the past 36 months.**

| | | **Name all entities with whom you have this relationship or indicate none (add rows as needed)** | **Specifications/Comments (e.g., if payments were made to you or to your institution)** |
|---|---|---|---|
| **Time frame: Since the initial planning of the work** | | | |
| 1 | All support for the present manuscript (e.g., funding, provision of study materials, medical writing, article processing charges, etc.) **No time limit for this item.** | None | |
| **Time frame: past 36 months** | | | |
| 2 | Grants or contracts from any entity (if not indicated in item #1 above). | None | |
| 3 | Royalties or licenses | None | |
| 4 | Consulting fees | None | |

| | | | |
|---|---|---|---|
| 5 | Payment or honoraria for lectures, presentations, speakers bureaus, manuscript writing or educational events | Honoraria for presentations | Sumitomo Dainippon Pharma Co., Ltd.<br>Payments were made to me. |
| | | Honoraria for a presentation | Astellas Pharma Inc.<br>Payments were made to me. |
| | | Honoraria for a presentation | FUJIFILM Toyama Chemical Co., Ltd.<br>Payments were made to me. |
| | | Honoraria for a presentation | MSD Co., Ltd.<br>Payments were made to me. |
| | | Honoraria for a presentation | Becton Dickinson Co., Ltd.<br>Payments were made to me. |
| 6 | Payment for expert testimony | None | |
| 7 | Support for attending meetings and/or travel | None | |
| 8 | Patents planned, issued or pending | None | |
| 9 | Participation on a Data Safety Monitoring Board or Advisory Board | None | |
| 10 | Leadership or fiduciary role in other board, society, committee or advocacy group, paid or unpaid | None | |
| 11 | Stock or stock options | None | |
| 12 | Receipt of equipment, materials, drugs, medical writing, gifts or other services | None | |
| 13 | Other financial or non-financial interests | None | |

**Please place an "X" next to the following statement to indicate your agreement:**

**_ X __ I certify that I have answered every question and have not altered the wording of any of the questions on this form.**

### ICMJE DISCLOSURE FORM

**Date: May 28, 2022**
**Your Name: Akitsugu Furumoto**
**Manuscript Title: Decreased hospitalizations and deaths from community-acquired pneumonia coincided with rising public awareness of personal precautions before the governmental containment and closure policy: A nationwide observational study in Japan**
**Manuscript number (if known):_______________________________________________________________**

**In the interest of transparency, we ask you to disclose all relationships/activities/interests listed below that are related to the content of your manuscript. "Related" means any relation with for-profit or not-for-profit third parties whose interests may be affected by the content of the manuscript. Disclosure represents a commitment to transparency and does not necessarily indicate a bias. If you are in doubt about whether to list a relationship/activity/interest, it is preferable that you do so.**

**The following questions apply to the author's relationships/activities/interests as they relate to the current manuscript only.**

**The author's relationships/activities/interests should be defined broadly. For example, if your manuscript pertains to the epidemiology of hypertension, you should declare all relationships with manufacturers of antihypertensive medication, even if that medication is not mentioned in the manuscript.**

**In item #1 below, report all support for the work reported in this manuscript without time limit. For all other items, the time frame for disclosure is the past 36 months.**

| | | **Name all entities with whom you have this relationship or indicate none (add rows as needed)** | **Specifications/Comments (e.g., if payments were made to you or to your institution)** |
|---|---|---|---|
| | | **Time frame: Since the initial planning of the work** | |
| 1 | All support for the present manuscript (e.g., funding, provision of study materials, medical writing, article processing charges, etc.) **No time limit for this item.** | None | |
| | | **Time frame: past 36 months** | |
| 2 | Grants or contracts from any entity (if not indicated in item #1 above). | None | |
| 3 | Royalties or licenses | None | |
| 4 | Consulting fees | None | |

| | | | |
|---|---|---|---|
| 5 | Payment or honoraria for lectures, presentations, speakers bureaus, manuscript writing or educational events | Honoraria for a presentation | Takeda Pharmaceutical company<br>Honoraria was sent to me. |
| | | Honoraria for a presentation | Sasebo medical association<br>Honoraria was sent to me. |
| | | Honoraria for presentation | Social welfare corporation Shirayuki Kai<br>Honoraria was sent to me. |
| | | Payment for a speaker | Nikkei Radio Broadcasting Corporation<br>Payment was sent to me. |
| | | Honoraria for presentations. | Nagasaki disaster rehabilitation council<br>Honoraria was sent to me. |
| | | Honoraria for a presentation | Nakano medical association<br>Honoraria was sent to me. |
| | | Honoraria for a presentation | Japan Organization of Occupational Health and Safety Nagasaki Rosai Hospital<br>Honoraria was sent to me. |
| | | Honoraria for a presentation | Nagasaki Prefecture government, Division of public health medical policy<br>Honoraria was sent to me. |
| | | Honoraria for a presentation | Sasebo City Office , division of public health and welfare<br>Honoraria was sent to me. |
| | | Honoraria for lectures | Nagasaki Nursing Association<br>Honoraria was sent to me. |
| | | Honoraria for manuscript writing | Nagasaki city medical association<br>Honoraria was sent to me. |
| 6 | Payment for expert testimony | None | |
| 7 | Support for attending meetings and/or travel | None | |
| 8 | Patents planned, issued or pending | None | |
| 9 | Participation on a Data Safety Monitoring Board or Advisory Board | None | |
| 10 | Leadership or fiduciary role in other board, society, committee or advocacy group, paid or unpaid | None | |
| 11 | Stock or stock options | None | |
| 12 | Receipt of equipment, materials, drugs, medical writing, gifts or other services | None | |

| | | |
|---|---|---|
| 13 | Other financial or non-financial interests | None |

**Please place an "X" next to the following statement to indicate your agreement:**

**_ X __ I certify that I have answered every question and have not altered the wording of any of the questions on this form.**

#### ICMJE DISCLOSURE FORM

**Date: May 26, 2022**
**Your Name: Katsunori Yanagihara**
**Manuscript Title: Decreased hospitalizations and deaths from community-acquired pneumonia coincided with rising public awareness of personal precautions before the governmental containment and closure policy: A nationwide observational study in Japan**
**Manuscript number (if known):_______________________________________________________________**

**In the interest of transparency, we ask you to disclose all relationships/activities/interests listed below that are related to the content of your manuscript. "Related" means any relation with for-profit or not-for-profit third parties whose interests may be affected by the content of the manuscript. Disclosure represents a commitment to transparency and does not necessarily indicate a bias. If you are in doubt about whether to list a relationship/activity/interest, it is preferable that you do so.**

**The following questions apply to the author's relationships/activities/interests as they relate to the current manuscript only.**

**The author's relationships/activities/interests should be defined broadly. For example, if your manuscript pertains to the epidemiology of hypertension, you should declare all relationships with manufacturers of antihypertensive medication, even if that medication is not mentioned in the manuscript.**

**In item #1 below, report all support for the work reported in this manuscript without time limit. For all other items, the time frame for disclosure is the past 36 months.**

| | | **Name all entities with whom you have this relationship or indicate none (add rows as needed)** | **Specifications/Comments (e.g., if payments were made to you or to your institution)** |
|---|---|---|---|
| **Time frame: Since the initial planning of the work** | | | |
| 1 | All support for the present manuscript (e.g., funding, provision of study materials, medical writing, article processing charges, etc.) **No time limit for this item.** | None | |
| **Time frame: past 36 months** | | | |
| 2 | Grants or contracts from any entity (if not indicated in item #1 above). | None | |
| 3 | Royalties or licenses | None | |
| 4 | Consulting fees | None | |

| | | |
|---|---|---|
| 5 | Payment or honoraria for lectures, presentations, speakers bureaus, manuscript writing or educational events | None |
| 6 | Payment for expert testimony | None |
| 7 | Support for attending meetings and/or travel | None |
| 8 | Patents planned, issued or pending | None |
| 9 | Participation on a Data Safety Monitoring Board or Advisory Board | None |
| 10 | Leadership or fiduciary role in other board, society, committee or advocacy group, paid or unpaid | None |
| 11 | Stock or stock options | None |
| 12 | Receipt of equipment, materials, drugs, medical writing, gifts or other services | None |
| 13 | Other financial or non-financial interests | None |

**Please place an "X" next to the following statement to indicate your agreement:**

**_ X __ I certify that I have answered every question and have not altered the wording of any of the questions on this form.**

**ICMJE DISCLOSURE FORM**

**Date: May27, 2022**
**Your Name: Hiroshi Mukae**
**Manuscript Title: Decreased hospitalizations and deaths from community-acquired pneumonia coincided with rising public awareness of personal precautions before the governmental containment and closure policy: A nationwide observational study in Japan**
**Manuscript number (if known):________________________________________________________________**

**In the interest of transparency, we ask you to disclose all relationships/activities/interests listed below that are related to the content of your manuscript. "Related" means any relation with for-profit or not-for-profit third parties whose interests may be affected by the content of the manuscript. Disclosure represents a commitment to transparency and does not necessarily indicate a bias. If you are in doubt about whether to list a relationship/activity/interest, it is preferable that you do so.**

**The following questions apply to the author's relationships/activities/interests as they relate to the current manuscript only.**

**The author's relationships/activities/interests should be defined broadly. For example, if your manuscript pertains to the epidemiology of hypertension, you should declare all relationships with manufacturers of antihypertensive medication, even if that medication is not mentioned in the manuscript.**

**In item #1 below, report all support for the work reported in this manuscript without time limit. For all other items, the time frame for disclosure is the past 36 months.**

| | | **Name all entities with whom you have this relationship or indicate none (add rows as needed)** | **Specifications/Comments (e.g., if payments were made to you or to your institution)** |
|---|---|---|---|
| **Time frame: Since the initial planning of the work** | | | |
| 1 | All support for the present manuscript (e.g., funding, provision of study materials, medical writing, article processing charges, etc.) **No time limit for this item.** | None | |
| **Time frame: past 36 months** | | | |
| 2 | Grants or contracts from any entity (if not indicated in item #1 above). | Grants or contracts | Nippon Boehringer Ingelheim Co ., Ltd. |
| | | Grants or contracts | Otsuka Pharmaceutical Co., Ltd. |
| | | Grants or contracts | SHIONOGI Co., Ltd. |
| | | Grants or contracts | Kyorin Pharmaceutical Co., Ltd. |
| | | Grants or contracts | Taisho Pharma Co., Ltd. |
| 3 | Royalties or licenses | None | |

| | | | |
|---|---|---|---|
| 4 | Consulting fees | None | |
| 5 | Payment or honoraria for lectures, presentations, speakers bureaus, manuscript writing or educational events | Payment or honoraria for lectures, presentations | Taiho Pharmaceutical Co., Ltd. |
| | | Payment or honoraria for lectures, presentations | Bristol-Myers Squibb |
| | | Payment or honoraria for lectures, presentations | Chugai Pharmaceutical Co., Ltd. |
| | | Payment or honoraria for lectures, presentations | Janssen Pharmaceutical K.K. |
| | | Payment or honoraria for lectures, presentations | Asahi Kasei Pharma Corporation |
| | | Payment or honoraria for lectures, presentations | Sumitomo Pharma Co., Ltd. |
| | | Payment or honoraria for lectures, presentations | Kyorin Pharmaceutical Co., Ltd. |
| | | Payment or honoraria for lectures, presentations | Taisho Pharma Co., Ltd. |
| | | Payment or honoraria for lectures, presentations | AstraZeneca K.K. |
| | | Payment or honoraria for lectures, presentations | Astellas Pharma Inc. |
| | | Payment or honoraria for lectures, presentations | SHIONOGI Co., Ltd. |
| | | Payment or honoraria for lectures, presentations | Daiichi Sankyo Co., Ltd. |
| | | Payment or honoraria for lectures, presentations | Pfizer Inc. |
| | | Payment or honoraria for lectures, presentations | MSD Co., Ltd |
| | | Payment or honoraria for lectures, presentations | Nippon Boehringer Ingelheim Co., Ltd. |
| | | Payment or honoraria for lectures, presentations | FUJIFILM Toyama Chemical Co., Ltd. |
| | | Payment or honoraria for lectures, presentations | Miyarisan Pharmaceutical Co., Ltd. |
| | | Payment or honoraria for lectures, presentations | Abbvie GK |
| | | Payment or honoraria for lectures, presentations | Novartis Pharma K.K |
| | | Payment or honoraria for lectures, presentations | Fisher & Paykel Healthcare Limited. |
| | | Payment or honoraria for lectures, presentations | Insmed Incorporated |
| | | Payment or honoraria for lectures, presentations | Sanofi K.K. |
| | | Payment or honoraria for lectures, presentations | Gilead Sciences Inc. |

| | | | |
|---|---|---|---|
| | | Payment or honoraria for lectures, presentations | KYOKUTO PHARMACEUTICAL INDUSTRIAL CO., LTD |
| | | Payment or honoraria for lectures, presentations | TAUNS Laboratories, Inc. |
| | | Payment or honoraria for lectures, presentations | Takeda Pharmaceutical Company Limited. |
| | | Payment or honoraria for lectures, presentations | bioMérieux Japan Ltd. |
| | | Payment or honoraria for lectures, presentations | Merck Biopharma Co., Ltd. |
| | | Payment or honoraria for lectures, presentations | Eli Lilly Japan K.K. |
| 6 | Payment for expert testimony | Payment for expert testimony | Janssen Pharmaceutical K.K. |
| | | Payment for expert testimony | Kyorin Pharmaceutical Co., Ltd. |
| | | Payment for expert testimony | SHIONOGI Co., Ltd. |
| | | Payment for expert testimony | Pfizer Inc. |
| | | Payment for expert testimony | MSD Co., Ltd |
| | | Payment for expert testimony | GlaxoSmithKline K.K. |
| | | Payment for expert testimony | Gilead Sciences Inc. |
| 7 | Support for attending meetings and/or travel | None | |
| 8 | Patents planned, issued or pending | None | |
| 9 | Participation on a Data Safety Monitoring Board or Advisory Board | None | |
| 10 | Leadership or fiduciary role in other board, society, committee or advocacy group, paid or unpaid | None | |
| 11 | Stock or stock options | None | |
| 12 | Receipt of equipment, materials, drugs, medical writing, gifts or other services | None | |
| 13 | Other financial or non-financial interests | None | |

**Please place an “X” next to the following statement to indicate your agreement:**

**_ X __ I certify that I have answered every question and have not altered the wording of any of the questions on this form.**

#### ICMJE DISCLOSURE FORM

**Date: May 26, 2022**
**Your Name: Kiyohide Fushimi**
**Manuscript Title: Decreased hospitalizations and deaths from community-acquired pneumonia coincided with rising public awareness of personal precautions before the governmental containment and closure policy: A nationwide observational study in Japan**
**Manuscript number (if known):________________________________________________________________**

**In the interest of transparency, we ask you to disclose all relationships/activities/interests listed below that are related to the content of your manuscript. "Related" means any relation with for-profit or not-for-profit third parties whose interests may be affected by the content of the manuscript. Disclosure represents a commitment to transparency and does not necessarily indicate a bias. If you are in doubt about whether to list a relationship/activity/interest, it is preferable that you do so.**

**The following questions apply to the author's relationships/activities/interests as they relate to the current manuscript only.**

**The author's relationships/activities/interests should be defined broadly. For example, if your manuscript pertains to the epidemiology of hypertension, you should declare all relationships with manufacturers of antihypertensive medication, even if that medication is not mentioned in the manuscript.**

**In item #1 below, report all support for the work reported in this manuscript without time limit. For all other items, the time frame for disclosure is the past 36 months.**

| | | **Name all entities with whom you have this relationship or indicate none (add rows as needed)** | **Specifications/Comments (e.g., if payments were made to you or to your institution)** |
|---|---|---|---|
| | | **Time frame: Since the initial planning of the work** | |
| 1 | All support for the present manuscript (e.g., funding, provision of study materials, medical writing, article processing charges, etc.) **No time limit for this item.** | None | |
| | | **Time frame: past 36 months** | |
| 2 | Grants or contracts from any entity (if not indicated in item #1 above). | None | |
| 3 | Royalties or licenses | None | |
| 4 | Consulting fees | None | |

| | | |
|---|---|---|
| 5 | Payment or honoraria for lectures, presentations, speakers bureaus, manuscript writing or educational events | None |
| 6 | Payment for expert testimony | None |
| 7 | Support for attending meetings and/or travel | None |
| 8 | Patents planned, issued or pending | None |
| 9 | Participation on a Data Safety Monitoring Board or Advisory Board | None |
| 10 | Leadership or fiduciary role in other board, society, committee or advocacy group, paid or unpaid | None |
| 11 | Stock or stock options | None |
| 12 | Receipt of equipment, materials, drugs, medical writing, gifts or other services | None |
| 13 | Other financial or non-financial interests | None |

**Please place an "X" next to the following statement to indicate your agreement:**

**_ X __ I certify that I have answered every question and have not altered the wording of any of the questions on this form.**

### ICMJE DISCLOSURE FORM

**Date: May 29, 2022**
**Your Name: Koichi Izumikawa**
**Manuscript Title: Decreased hospitalizations and deaths from community-acquired pneumonia coincided with rising public awareness of personal precautions before the governmental containment and closure policy: A nationwide observational study in Japan**
**Manuscript number (if known):_______________________________________________________________**

**In the interest of transparency, we ask you to disclose all relationships/activities/interests listed below that are related to the content of your manuscript. "Related" means any relation with for-profit or not-for-profit third parties whose interests may be affected by the content of the manuscript. Disclosure represents a commitment to transparency and does not necessarily indicate a bias. If you are in doubt about whether to list a relationship/activity/interest, it is preferable that you do so.**

**The following questions apply to the author's relationships/activities/interests as they relate to the <u>current manuscript only</u>.**

**The author's relationships/activities/interests should be <u>defined broadly</u>. For example, if your manuscript pertains to the epidemiology of hypertension, you should declare all relationships with manufacturers of antihypertensive medication, even if that medication is not mentioned in the manuscript.**

**In item #1 below, report all support for the work reported in this manuscript without time limit. For all other items, the time frame for disclosure is the past 36 months.**

| | | **Name all entities with whom you have this relationship or indicate none (add rows as needed)** | **Specifications/Comments (e.g., if payments were made to you or to your institution)** |
|---|---|---|---|
| | | **Time frame: Since the initial planning of the work** | |
| 1 | All support for the present manuscript (e.g., funding, provision of study materials, medical writing, article processing charges, etc.) **No time limit for this item.** | None | |
| | | **Time frame: past 36 months** | |
| 2 | Grants or contracts from any entity (if not indicated in item #1 above). | None | |
| 3 | Royalties or licenses | None | |
| 4 | Consulting fees | Asahi Kasei Pharma | payments were made to myself |

| | | | |
|---|---|---|---|
| | | Corporation | |
| 5 | Payment or honoraria for lectures, presentations, speakers bureaus, manuscript writing or educational events | Merck & Co., Inc. | honoraria were made to myself |
| | | Pfizer Inc. | honoraria were made to myself |
| | | Astellas Pharma Inc. | honoraria were made to myself |
| | | Asahi Kasei Pharma Corporation | honoraria were made to myself |
| | | Sumitomo Pharma Co., Ltd | honoraria were made to myself |
| | | KYORIN Pharmaceutical Co., Ltd | honoraria were made to myself |
| | | DAIICHI SANKYO COMPANY, LIMITED | honoraria were made to myself |
| | | FUJIFILM Toyama Chemical Co., Ltd. | honoraria were made to myself |
| | | Maruishi Pharmaceutical. Co., Ltd. | honoraria were made to myself |
| | | Gilead Sciences, Inc. | honoraria were made to myself |
| | | Taisho Pharmaceutical Co., Ltd. | honoraria were made to myself |
| | | SHIONOGI & Co., Ltd. | honoraria were made to myself |
| | | AstraZeneca K.K. | honoraria were made to myself |
| | | Kao Corporation. | honoraria were made to myself |
| 6 | Payment for expert testimony | None | |
| 7 | Support for attending meetings and/or travel | None | |
| 8 | Patents planned, issued or pending | None | |
| 9 | Participation on a Data Safety Monitoring Board or Advisory Board | None | |
| 10 | Leadership or fiduciary role in other board, society, committee or advocacy group, paid or unpaid | None | |
| 11 | Stock or stock options | None | |
| 12 | Receipt of equipment, materials, drugs, medical writing, gifts or other services | None | |
| 13 | Other financial or non-financial interests | None | |

**Please place an "X" next to the following statement to indicate your agreement:**

**_ X __ I certify that I have answered every question and have not altered the wording of any of the questions on this form.**
